# Treatment Approaches for Problematic Usage of the Internet (PUI): A Dual-Level Meta-Analysis of Meta-Analyses and Randomized Controlled Trials

**DOI:** 10.1101/2025.10.02.25337159

**Authors:** Alireza Valyan, Fateme Sadat Abolghasemi, Sophia Achab, Mitra Ashrafi, Alexander M. Baldacchino, Zsolt Demetrovics, Naomi A. Fineberg, Parastoo Ghorbani, Yasser Khazaal, Kristiana Siste, Anise M.S. Wu, Mehran Zare-Bidokey, Dan J. Stein, Marc N. Potenza, Hamed Ekhtiari

**Author notes:** **Corresponding Author:** Hamed Ekhtiari, MD, PhD Department of Psychiatry and Behavioral Sciences, University of Minnesota.

## Abstract

**Background:** Problematic usage of the internet (PUI) is a growing public health concern, with a rapidly expanding literature of primary trials and meta-analyses on treatment approaches. However, this evidence base remains fragmented due to differences in definitions, assessment tools, and methodological approaches, which limit its applicability for clinicians, policymakers, and researchers. To address this gap, we conducted a dual-level meta-analysis: first, a systematic review and meta-analysis of existing meta-analyses; second, a secondary meta-analysis of randomized controlled trials extracted from these studies.

**Methods:** Following PRISMA guidelines, we systematically searched PubMed between 2013 and 2024 for meta-analyses assessing treatments for a range of PUI-related conditions, including internet addiction and problems related to online gaming, online pornography, online shopping, social media use, and smartphone use. Eligible studies evaluated behavioral, pharmacological, neuromodulatory, or physical exercise interventions. Studies were excluded if they were not meta-analyses, did not focus on PUI, or lacked sufficient data. Methodological quality was assessed using AMSTAR 2.0 and Cochrane Risk of Bias 2.0 tool for the meta-analyses and RCTs, respectively. A random-effects model was applied to analyze pooled effect sizes, accounting for heterogeneity and publication bias. To enhance rigor, we conducted a secondary meta-analysis of randomized controlled trials (RCTs) extracted from these meta-analyses. This study was registered with the OSF (Open Science Framework) at https://osf.io/8uc2w.

**Findings:** Among 402 identified records, 329 were screened, and 20 meta-analyses met the inclusion criteria, comprising 35 units of analysis. Results of the meta-annalysis of meta-analyses confirmed the overall impact of PUI treatments (Standardized Mean Difference [SMD] = −1.41, 95% CI: [−1.54; −1.18], p < 0.0001), with variations across PUI dimensions, treatment categories, and age groups. In the second phase, from the identified 386 unique studies included in the eligible meta-analyses, 44 RCTs were identified and included for the secondary meta-analysis. Behavioral interventions, including CBT and its variants, were the most commonly used approaches (91 units of analysis in 35 RCTs) and showed a significant impact (SMD = –1.91; 95% CI: [–2.22, –1.60]). Neuromodulatory interventions showed the largest effect size (SMD = –3.14; 95% CI: [–3.14, –3.14]) but were underexplored (six units in two RCTs). Pharmacological interventions were also effective (SMD = –1.34; 95% CI: [–1.51, –1.18]) across 14 units in five RCTs. Physical exercise was effective with a lower effect size (SMD = –1.15; 95% CI: [– 1.77, –0.53]) based on eight units in three RCTs. Furthermore, serious concerns were detected with regard to the quality of studies, risks of bias, and methodological rigor at both levels.

**Interpretation:** This study provides an overview of the available evidence for PUI treatments and their observed outcomes. Behavioral interventions, particularly CBT, are supported by a substantial body of evidence showing positive results, while pharmacological and neuromodulatory treatments appear promising based on a smaller number of studies. Advancing the field will require RCTs with greater methodological rigor, standardized assessment tools, longitudinal designs, and culturally adapted interventions to improve the accessibility and impact of these treatments.

## 1 Introduction and Background

Problematic usage of the internet (PUI) has emerged as a significant public health concern (Achab et al. 2015; Fineberg et al. 2025). The World Health Organization (WHO) has acknowledged that excessive use of the internet and other digital electronic devices can lead to negative health consequences, with the issue reaching the magnitude of a significant public health concern in multiple global jurisdictions (World Health Organization, 2015, 2018). Prevalence estimates of PUI vary across different populations (Rochat et al. 2021; Achab and Billieux 2022; Achab 2023) and regions (Fernández et al. 2023; Fernández et al. 2025). Studies have reported prevalence estimates ranging from 2% to 35% (see (Gjoneska et al. 2025) for a review), indicating considerable heterogeneity due to factors such as lack of standardization (Carragher et al. 2022b) or uniform definition and lack of national epidemiological studies (Long et al. 2022; Kathiresan and Bhargava 2021) or heterogeneity of the assessment methods and diagnostic criteria (Rochat et al. 2025). Young people and individuals in low- and middle-income countries may be at elevated risk of PUI (Kokka et al. 2021; Dadi et al. 2024). Negative consequences associated with PUI are extensive, including impaired academic performance (Abdullahi et al. 2024); poor mental health, such as depression and anxiety (Jia et al. 2024; Cai et al. 2023); impaired social relationships (Cai et al. 2023; Rizzo and Alparone 2024); and other health concerns such as obesity (Yıldız et al. 2024). These consequences affect the overall quality of life and well-being. Underlying cognitive and behavioral mechanisms of PUI may be diverse among affected individuals and require targeted psychotherapeutic approaches (Fernández et al. 2022).

The global scale of PUI has led to an intensified focus on understanding this phenomenon (Cekic et al. 2024) and developing effective interventions (Fineberg et al. 2025). Given the widespread prevalence and significant impact of PUI, it is imperative to develop effective interventions and public health strategies either to prevent (Saletti et al. 2021) or mitigate (Chadha et al. 2024) its effects and promote healthier patterns of internet usage.

While numerous intervention strategies have been proposed and evaluated for PUI (Yang et al. 2024), findings across studies vary, and there is no uniform approach regarding therapeutic strategies (Dewi et al. 2024). Interventions such as cognitive behavioral therapy (CBT) (Furukawa et al. 2024) and pharmacotherapies (Łukawski et al. 2019) have been identified as potential treatments for PUI.

On the other hand, although each of these methods addresses specific aspects of PUI, the diversity of findings across studies has made it difficult to identify definitive, evidence-based treatments (Ayub et al. 2023). This challenge is compounded by the heterogeneous nature of PUI, which encompasses a spectrum of behaviors and diagnoses that often vary based on factors including the specific dimension of internet use (Eşkisu et al. 2024), As a result, understanding which interventions are most effective for sub-dimensions of PUI (problematic online gaming, online shopping, etc.) is not well-established.

The existing literature on PUI treatment has benefited from several systematic reviews and meta-analyses, which have provided insights into the impacts of various interventions. However, these studies are limited in scope and often fail to capture the full complexity of the issue. Many meta-analyses have focused on only one or a few dimensions of PUI, such as online gaming or social media use, and have not addressed its broader spectrum. Additionally, the divergence in PUI terminologies (Zare-Bidoky et al. 2025) and assessment tools (Schlossarek et al. 2024; Browne et al. 2021) across studies has hindered comparisons and synthesis of findings. While some reviews have examined specific interventions (e.g., (Liu et al. 2019))) or populations (e.g., (Malinauskas and Malinauskiene 2019)), few have attempted to systematically synthesize outcomes across multiple treatment modalities (e.g., (Jiang et al. 2023)); (Zhang et al. 2024)). Furthermore, the methodological quality of included studies has been inconsistent, with many relying on non-randomized designs or small sample sizes, limiting the generalizability of their findings. Our study addresses these gaps by integrating multiple PUI dimensions under a single umbrella term and conducting a dual-level meta-analysis to provide a comprehensive evaluation of treatment efficacy.

A key challenge in the literature is the lack of standardized tools for assessing PUI and its treatment outcomes (Fernández et al. 2022; Carragher et al. 2022a). This diversity of outcome measures has resulted in considerable variability in reported findings, complicating efforts to compare interventions—an issue our study aims to address by synthesizing findings across multiple treatment modalities.

Given the growing number of studies on PUI interventions, there has yet to be a comprehensive synthesis of existing meta-analyses. To address this gap, we implemented a novel dual-level approach, consisting of two stages. In the first stage, we compiled and analyzed published meta-analyses to capture the broader landscape of evidence—a step that has some precedent in mental health research on topics such as positive psychology (Carr et al. 2024) (Carr et al., 2024), attention-deficit/hyperactivity disorder (Betancourt et al. 2024) (Betancourt et al., 2024), anxiety (Nair et al. 2024), and substance use disorders (Mehta et al. 2024). In the second stage, we conducted a focused meta-analysis of randomized controlled trials (RCTs) identified through this review, emphasizing higher-quality data. The analytic focus in Phase 2 was exclusively on RCTs, as they represent the highest standard of evidence for evaluating treatment effectiveness and are widely regarded as the gold standard for minimizing bias and controlling for confounding factors (Cuello-Garcia et al. 2022).

This dual-level design is, to our knowledge, the first of its kind in PUI research and aims to provide a clearer picture of the effectiveness of current treatment strategies, offering evidence to support clinicians and researchers in guiding intervention choices.

## 2 Methods

### Study Design

“This study followed a two-phase systematic approach to evaluate treatment approaches for PUI. In Phase 1, we conducted a systematic review and meta-analysis of published meta-analyses (Eisenhauer 2021). In Phase 2, we identified and analyzed individual RCTs extracted from the included meta-analyses in the first phase. The study selection process at both levels adhered to the PRISMA guidelines (Moher et al. 2009). A detailed flow diagram illustrating the number of studies identified, screened, excluded, and included is provided in the Results section. The protocol for this study was pre-registered on the Open Science Framework (OSF) to ensure methodological transparency (accessible at: https://osf.io/8uc2w/).

### Search Strategy

A comprehensive search was conducted in PubMed to identify relevant meta-analyses addressing treatment interventions for PUI. Following previous studies regarding PUI nomenclature (Dahl and Bergmark 2020; Fernandes et al. 2019), the search targeted studies covering various dimensions of PUI, including general internet use, online gaming, online pornography, online shopping, social media use, and smartphone use. The search string, which included specific terms for each dimension along with Boolean operators, is provided in **Error! Reference source not found.**. Additionally, reference lists of included reviews and meta-analyses were manually screened to identify any additional relevant studies.

### Inclusion and Exclusion Criteria

The objective of this study was to systematically evaluate and synthesize evidence on treatments for PUI and its specific dimensions. Thus, the inclusion criteria for Phase 1 required that studies address PUI or its specific dimensions (e.g., relating to online gaming, social media, smartphone use, online shopping, or online pornography), evaluated treatment interventions specifically targeting PUI, were published in English, and were meta-analyses with sufficient data available for extraction and analysis. Studies were excluded if they did not focus on PUI or treatment interventions for PUI, were published in languages other than English, or were narrative reviews, scoping reviews, or systematic reviews without meta-analytic components or insufficient data for analysis.

In Phase 2, individual clinical trials from the selected meta-analyses were considered for inclusion. Eligible trials were randomized clinical trials that evaluated treatment interventions for PUI, were published in peer-reviewed journals, and were reported in English. Exclusion criteria included non-randomized or uncontrolled designs (e.g., observational or cross-sectional studies), studies lacking data on treatment interventions or treatment outcomes, and studies not published in peer-reviewed journals or not written in English. Additionally, studies addressing behaviors sometimes labeled as PUI but occurring offline—such as certain forms of shopping or pornography consumption—were excluded. This criterion was applied to ensure conceptual consistency, as mixing online and offline behaviors in the same analysis can obscure the interpretation of treatment effectiveness for truly online-related PUI.

The screening and inclusion of studies were independently conducted by three authors (P.G., F.S.A., and M.A.). Any conflicts arising during this process were resolved through group discussions led by A.V. and supervised by H.E.

### Data Extraction and Synthesis

For Phase 1, two of the co-authors (F.S.A. and P.G.) screened titles and abstracts, retrieved full texts, and applied the inclusion criteria. Any discrepancies were resolved by a third reviewer (A.V.). Detailed characteristics such as participant demographics, treatment categories, PUI dimensions, and effect sizes were extracted.

In Phase 2, studies were identified from the included meta-analyses of Phase 1. After removing duplications and applying the inclusion criteria, eligible trials were found. Data from these trials were extracted, including sample size, participant characteristics, PUI assessment tools, treatment modalities, control conditions, and outcomes. Data extraction was independently performed by three co-authors (P.G., F.S.A., and M.A.). Discrepancies were resolved in group discussions led by A.V. and under the supervision of H.E.

### Quality Assessment

To assess the methodological quality of systematic reviews and meta-analyses, the AMSTAR 2.0 checklist (Shea et al. 2017) was applied. For the clinical trials included in Phase 2, the Cochrane Risk of Bias 2.0 tool (Sterne et al. 2019) was used to evaluate study-level bias. The assessment of the risk of bias was conducted independently by three authors (P.G., F.S.A., and M.A.). Any disagreements were addressed in group discussions led by A.V. and under the supervision of H.E. The risk of publication bias was assessed visually using funnel plots and Egger’s regression test (Egger et al. 1997).

### Data Synthesis and Statistical Analyses

All statistical analyses were conducted using R 4.3.1 software. The standardized mean difference (SMD) was used as the measure of effect size. In studies reporting multiple outcomes or comparison groups, effect sizes were aggregated within a study using a weighted average so that each study contributed a single, independent estimate. A random-effects model was used to account for the between-study heterogeneity, estimated using restricted maximum likelihood (Viechtbauer 2010). The I² statistic was used to quantify the heterogeneity, with values above 50% indicating substantial heterogeneity. When I² exceeded 50%, outlier and influence analyses were performed to examine the sources of heterogeneity. Additionally, the H² statistic was calculated to assess the ratio of observed variation to expected variance due to sampling error, and the variance (τ²) and its standard deviation (τ) were determined to quantify heterogeneity.

Subgroup analyses and meta-regression were performed to examine the impact of participant age group, cultural background (non-Eastern vs. Eastern), PUI scale category (symptoms vs. consumption), risk of bias (low, some concerns, and high), intervention categories, types, and control conditions on effect sizes.

## 3 Results

### Meta-Analysis of Meta-Analyses (Phase 1)

#### Characteristics of Included Meta-Analyses

In the first phase, a total of 20 meta-analyses were included for further analysis. The selection process, summarized in **Figure 1**-Top, began with identifying 402 systematic reviews and meta-analyses through a database search, supplemented by 31 studies from forward and backward citation tracking from the inception till the end of 2024. After removing 103 duplicates, 330 studies were sought for retrieval. Among these, one study could not be retrieved due to the unavailability of a full text, leaving 329 studies for screening.

**Figure 1:**
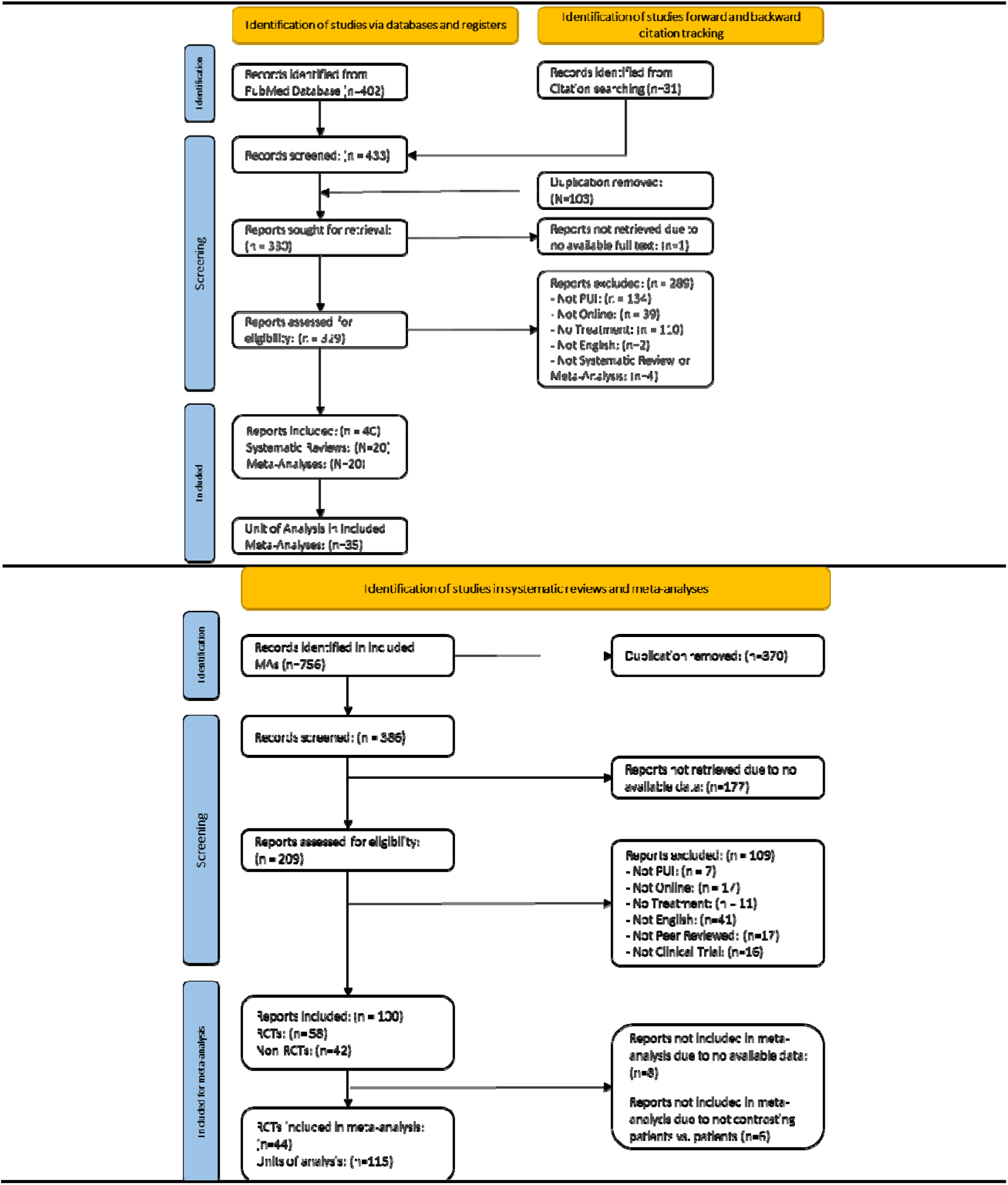
Overview of study selection processes in both phases. The top panel illustrates the PRISMA flow diagram used to identify eligible systematic reviews and meta-analyses on treatments for problematic usage of the internet (PUI). The bottom panel presents the PRISMA diagram for Phase 2, outlining the identification of randomized controlled trials (RCTs) extracted from the included meta-analyses.

During the screening process, 289 studies were excluded for various reasons: 134 did not focus on PUI, 39 were not related to online contexts, 110 did not involve treatment, one was not in English, and four were not systematic reviews or meta-analyses. This resulted in 40 eligible systematic reviews and meta-analyses for data extraction. Since our study focused exclusively on meta-analyses, the 20 systematic reviews without quantitative synthesis were not retained, leaving 20 meta-analyses for further analysis. These 20 meta-analyses comprised a total of 35 units of analysis, which were the focus of the subsequent evaluation. The complete list of screened systematic reviews and meta-analyses can be found on the project’s OSF page at https://osf.io/8uc2w/.

*The characteristics of the included meta-analyses are summarized in*

Table 1. These reviews represent diverse research efforts published between 2013 and 2024 (**Error! Reference source not found.**,b). These studies included 6 to 91 studies (average = 34) with an inclusion rate between 0.09% to 20.97% (average = 2.45%), reflecting differences in inclusion criteri and scope. Collectively, they included data from a total of 756 studies, representing 41,525 participants, with sample sizes ranging from 305 to 5,601 participants (average = 2146). Most included meta-analyse (13, 68%) combined the results for mixed participant age groups, with fewer meta-analyses focusing on adolescents (3), young adults (3), and children (1) (**Error! Reference source not found.**,c).

**Table 1:** Summary of the 20 Meta-Analyses on the Treatment of the Problematic Usage of the Internet (PUI). This table provides an overview of the 20 meta-analyses selected from an initial pool of 402 records. It summarizes key characteristics, including publication year, number of included studies (N), inclusion rate, total participants (K), targeted PUI dimensions, outcome measures, treatment categories, effect estimates, and assessed risk of bias. The table supports a comparative understanding of treatment approaches and outcome trends across different PUI dimensions.

| Study | Year | N | Inclusion Rate | K | Participants | PUI Dimension | Outcome Measures | Treatment Category | Effect | Study Quality |
| --- | --- | --- | --- | --- | --- | --- | --- | --- | --- | --- |
| (Andrade et al. 2022) | 2022 | 15 | 0.6% | 1,671 | Mixed | Internet Addiction | YIAS, CIAS, YDQ, PIUQ, TMDS, SI, BDQIA | Behavioural, Pharmacological, Physical, Neuromodulatory | -1.29 | Low |
| (Augner et al. 2022) | 2021 | 14 | 0.3% | 1,439 | Mixed | Internet Addiction, Smartphone | YIAS, CIAS, SAS, YDQ, MPIAS, APUIS, BSMAS, KSAPS, AIADS | Behavioural | -0.89 | Low |
| (Chang et al. 2022) | 2022 | 26 | 21.0% | 5,601 | Young Adults | Internet Addiction, Online Gaming | YIAS, CIAS, APUIS, CGAI, TMDS, IADSS | Behavioural, Pharmacological, Physical, Neuromodulatory | -1.39 | Critically Low |
| (Fendel et al. 2024) | 2024 | 39 | 1.1% | 1,549 | Mixed | Internet Addiction | YIAS, CIAS, SAS, YDQ, MPIAS, MPPUS, BSMAS, MPBACSAS, IOSRS, IAIUS K-SCALE, DPVQ, YGIAT, SMUQ, STSR, VGAS, PPCS, CPUI, BPS, SPAL, IUQ | Behavioural | -1.67 | Low |
| (Goslar et al. 2020) | 2020 | 91 | 1.2% | 2,427 | Mixed | Internet Addiction, Online Pornography, Online Shopping | YIAS, CIAS, YDQ, MPIAS, YBOCS-ICIUD, GAST, APUIS, IGT, KSAPS, PIUQ, IOSRS, AICA-S, PVGPS, IUT, DPVQ, SIUS, CCU, DSM5-IGD, YBOCS-SV, DQVMIA, AASEOM, IGD-C, GASA, APSOA | Behavioural, Pharmacological, Physical, Neuromodulatory | -1.27 | Low |
| (Jiang et al. 2023) | 2024 | 66 | 0.2% | 4,385 | Mixed | Internet Addiction | YIAS, YDQ, OGAS, CGAI, PIUQ, AICA-S | Behavioral, Physical, Neuromodulatory | -2.01 | Low |
| (Kim and Noh 2019) | 2019 | 11 | 0.3% | 658 | Mixed | Internet Addiction, Online Gaming | YIAS, CIAS, YDQ, APUIS, IGT, OGCAS, IOSRS, AICA-S, TMDS, IUT, BDQIA | Behavioral, Pharmacological | -0.79 | Critically Low |
| (Li et al. 2023) | 2023 | 12 | 0.3% | 861 | Adolescents | Smartphone | SAS, MPATS, MPIAS | Physical | -3.11 | Low |
| (Liu et al. 2017) | 2017 | 58 | 5.7% | 2,871 | Mixed | Internet Addiction | YIAS, YDQ, APUIS | Behavioral, Physical | -1.63 | Low |
| (Liu et al. 2019) | 2019 | 9 | 6.3% | 1,582 | Young Adults | Smartphone | SAS, MPATS, MPIAS | Physical | -1.30 | Low |
| (Malinauskas and Malinauskiene 2019) | 2019 | 6 | 0.4% | 305 | Adolescents | Internet Addiction, Smartphone | YIAS, BSMAS, PIUQ | Behavioral, Pharmacological | -0.68 | Critically Low |
| (Stevens et al. 2019) | 2018 | 13 | 0.8% | 580 | Mixed | Online Gaming | YIAS, CIAS, MTQ, IOSRS, AICA-S, IUT, RF, ATA, ACU, AEO | Behavioural | -0.92 | Critically Low |

Dual-Level Meta-Analysis of PUI Treatments
| Study | Year | N | Inclusion Rate | K | Participants | PUI Dimension | Outcome Measures | Treatment Category | Effect | Study Quality |
| --- | --- | --- | --- | --- | --- | --- | --- | --- | --- | --- |
| (Wang et al. 2024) | 2024 | 18 | 0.1% | 1,158 | Mixed | Internet Addiction | CIAS, GAST, GPUIS, APUIS, GAME-P, CIUS-P | Behavioural | -1.43 | Low |
| (Winkler et al. 2013) | 2013 | 16 | 2.4% | 670 | Mixed | Internet Addiction, Online Gaming, Online Pornography, Online Shopping | YIAS, CIAS, YDQ, MTS, IOSRS, IADSS, IUT, ATA, ATC, SOIAS | Behavioral, Pharmacological | -1.35 | Critically Low |
| **(Yan et al. 2024)** | 2024 | 15 | 0.5% | 760 | Young Adults | Internet Addiction | YIAS, CIAS, MPAI, MPATS | Physical | -1.25 | Low |
| (Yeun and Han 2016) | 2016 | 37 | 1.8% | 1,490 | Children | Internet Addiction | YIAS, CIAS, YDQ, IGAS | Behavioural | -1.19 | Low |
| (Zhang et al. 2024) | 2022 | 24 | 0.4% | 3,832 | Mixed | Internet addiction, Online Gaming, Online Pornography | YIAS, CIAS, YDQ | Behavioural, Pharmacological, Neuromodulatory | -1.90 | Low |
| (Zhang et al. 2022) | 2024 | 44 | 1.0% | 2,654 | Young Adults | Internet Addiction | YIAS, CIAS | Behavioural, Pharmacological, Behavioural | -1.70 | Low |
| (Zhou et al. 2024) | 2024 | 89 | 1.3% | 3,494 | Adolescents | Internet Addiction | YIAS, CIAS, MPIAS, PIUQ, IADSS, DSM5-IGD, IGD-C | Behavioural, Pharmacological, Neuromodulatory | -1.75 | Low |
| (Zhu et al. 2023) | 2023 | 57 | 1.5% | 3,538 | Mixed | Internet Addiction, Smartphone, Online Gaming | YIAS | Behavioural, Pharmacological, Neuromodulatory | -1.57 | Low |

*The included meta-analyses examined a wide array of PUI dimensions, including general internet addiction, online gaming, online pornography, smartphone use, social media use, and online shopping. The frequency of PUI dimensions analyzed across the included meta-analyses is shown in*

Figure 2-Left. Notably, internet addiction was the most frequently studied dimension, followed by problematic smartphone use and problematic online gaming. Interventions across the included meta-analyses were categorized into four main treatment types: behavioral, pharmacological, neuromodulatory, and physical exercise. Behavioral interventions were the most frequently evaluated, followed by pharmacological treatments. The frequency distribution of these treatment categories is presented in Figure 2-Right.

**Figure 2:**
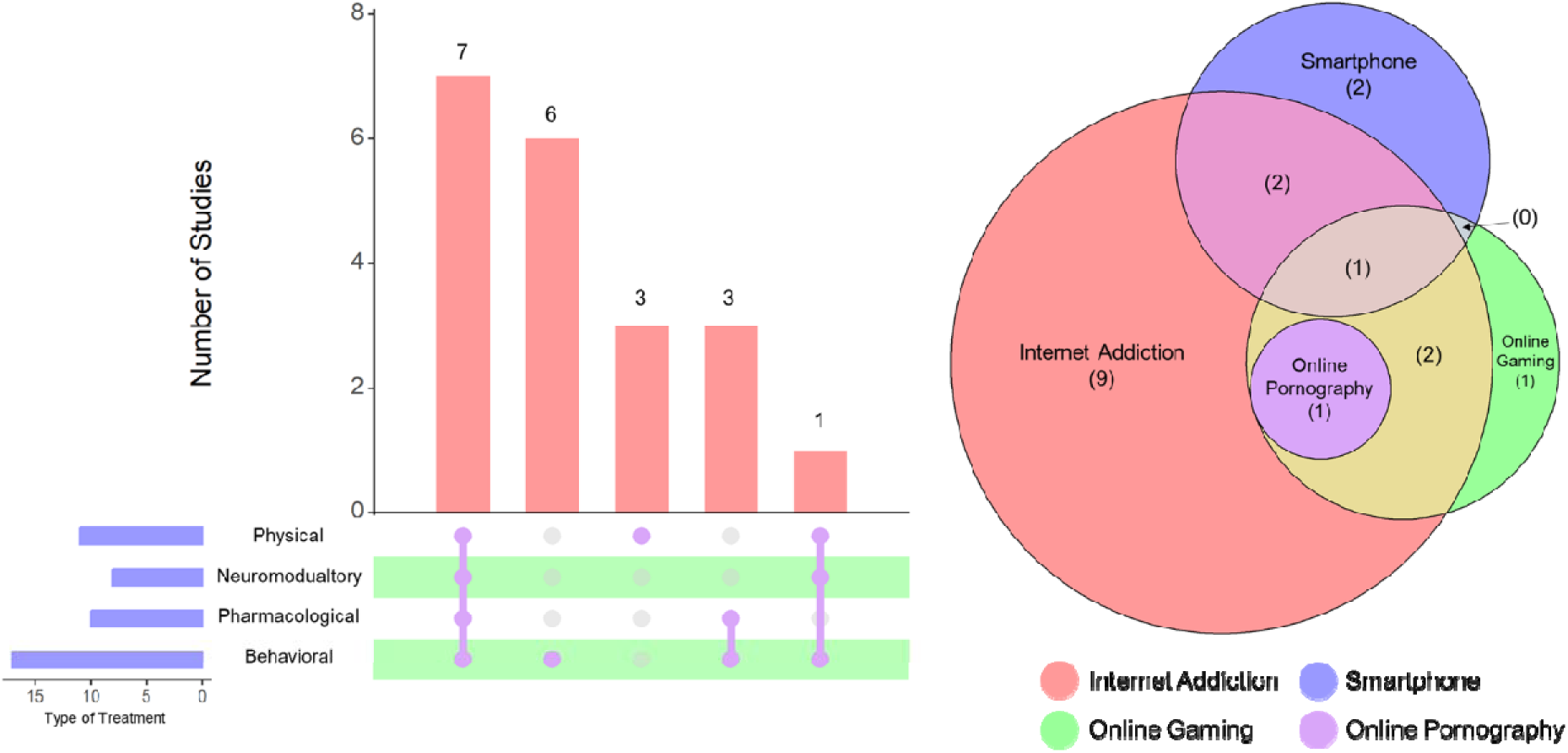
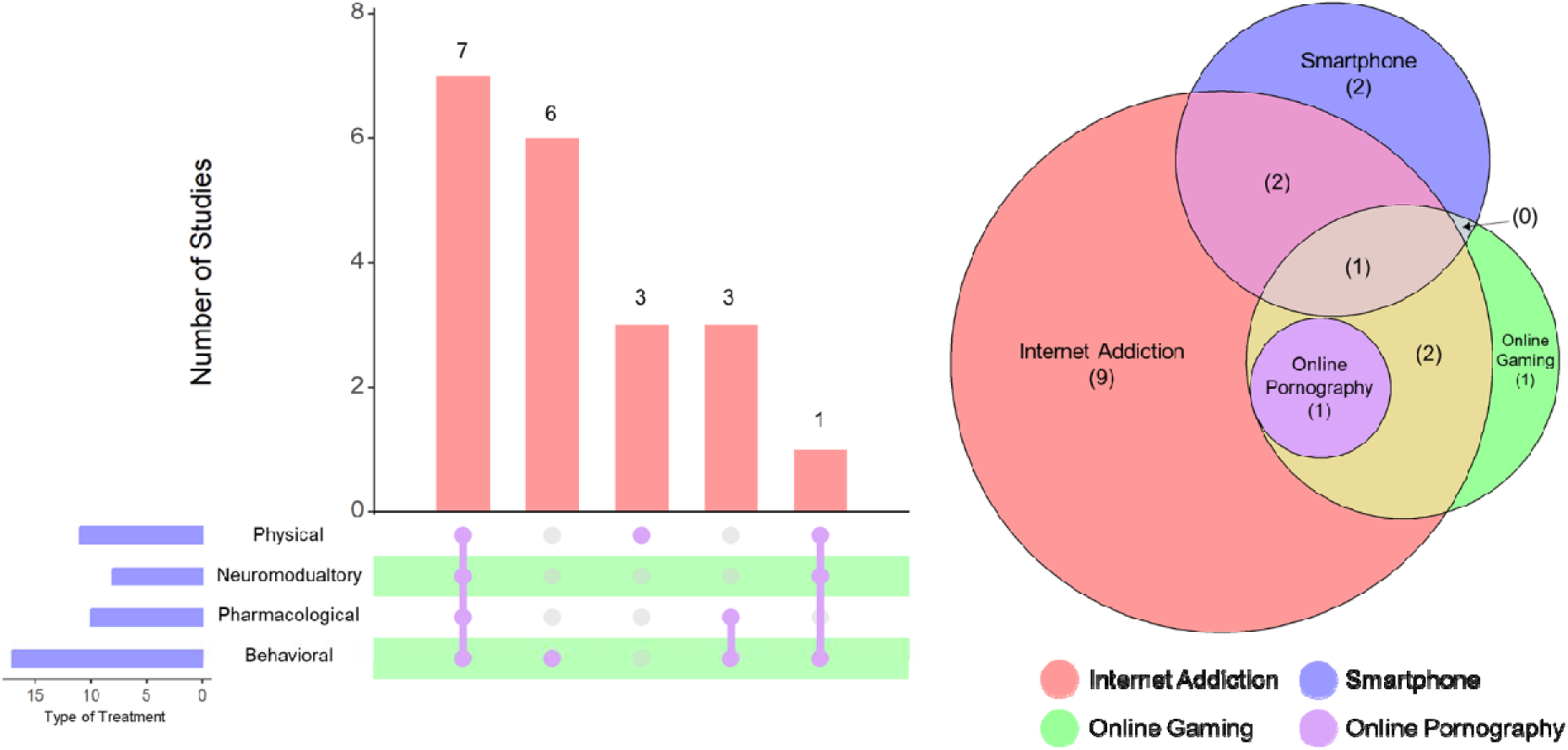
Overview of the scope and treatment focus of included meta-analyses. The left panel illustrates the types of treatments evaluated, including behavioral, pharmacological, neuromodulatory, and physical interventions, with attention to studies assessing combined approaches. The right panel shows the distribution of PUI dimensions addressed across the included meta-analyses, highlighting patterns in topic representation and overlap.

### Analytic Results of Meta-Analyses

The meta-analytical synthesis of the 35 units of analysis derived from the 20 included meta-analyses yielded a statistically significant pooled random-effects size of -1.42 (95% CI: [-1.63; -1.22], t=-14.26, p < 0.0001). The prediction interval ranged from [-2.48; -0.38], indicating the variability in potential true effect sizes across different contexts. The list of all units of analysis can be found in **Error!** Reference source not found..

Substantial heterogeneity was observed, with a tau-squared ( ^2^) value of 0.26 [0.15; 0.55] and an I-squared (I^2^) statistic of 83.4% (95% CI: [77.7%; 87.6%]), demonstrating high inconsistency across studies. Given the substantial heterogeneity (I^2^>50%), further exploration was warranted to identify it sources. While we considered calculating effect sizes separately for each PUI dimension, this approach was not feasible due to the inclusion of multiple meta-analyses that combined their effect sizes acros several PUI dimensions. Examples include (Zhu et al. 2023) (internet addiction, smartphone, and online gaming), (Zhang et al. 2022) (internet addiction, online gaming, and online pornography), (Malinauska and Malinauskiene 2019) (Internet Addiction and Smartphone), and (Chang et al. 2022) (Internet Addiction and Online Gaming). Nonetheless, calculating effect sizes separately for each PUI dimension did not affect heterogeneity significantly except for the case of problematic online gaming, where the I^2^ value decreased to 44%, but with no change in p-value of 0.18 (see Error! Reference source not found.-left)

Furthermore, most included meta-analyses utilized combined treatments. Examples include (Chang et al. 2022) (behavioral + pharmacological + neuromodulatory), (Goslar et al. 2020) (behavioral + pharmacological), (Jiang et al. 2023) (behavioral + physical), (Winkler et al. 2013) (behavioral + pharmacological), (Zhang et al. 2022) (behavioral + pharmacological + neuromodulatory), (Zhang et al. 2024) (behavioral + physical), (Zhou et al. 2024) (behavioral + physical + pharmacological + neuromodulatory), and (Zhu et al. 2023) (behavioral + pharmacological + neuromodulatory). Here, calculating effect sizes for each treatment category did not affect heterogeneity, except for studies using behavioral + physical treatments, where the I^2^ value dropped to 43%, but the p-value remained at 0.19.

Given these combinations in both PUI dimensions and treatment categories, the next step involved identifying potential outliers to better understand the sources of heterogeneity and their potential impact on the overall findings. Outliers were identified as meta-analyses whose confidence intervals did not overlap with the overall pooled estimate. Specifically, meta-analyses were considered outliers if their upper confidence limit was below the overall lower confidence limit or their lower confidence limit was above the overall upper confidence limit. After applying this criterion, eight outliers were detected and removed from the analysis. The updated analysis, which included the remaining 27 units of analysis, yielded a pooled random-effects size of -1.41 (95% CI: [-1.54; -1.29], t= −23.24, p < 0.0001), with a prediction interval of [-1.85; -0.97] (Figure 3). Notably, while the pooled effect size did not vary significantly after the removal of outliers, the heterogeneity statistics showed a dramatic reduction. The I^2^ value decreased from 84.1% to 51.0%, indicating a much more moderate level of heterogeneity among the remaining studies.

**Figure 3:**
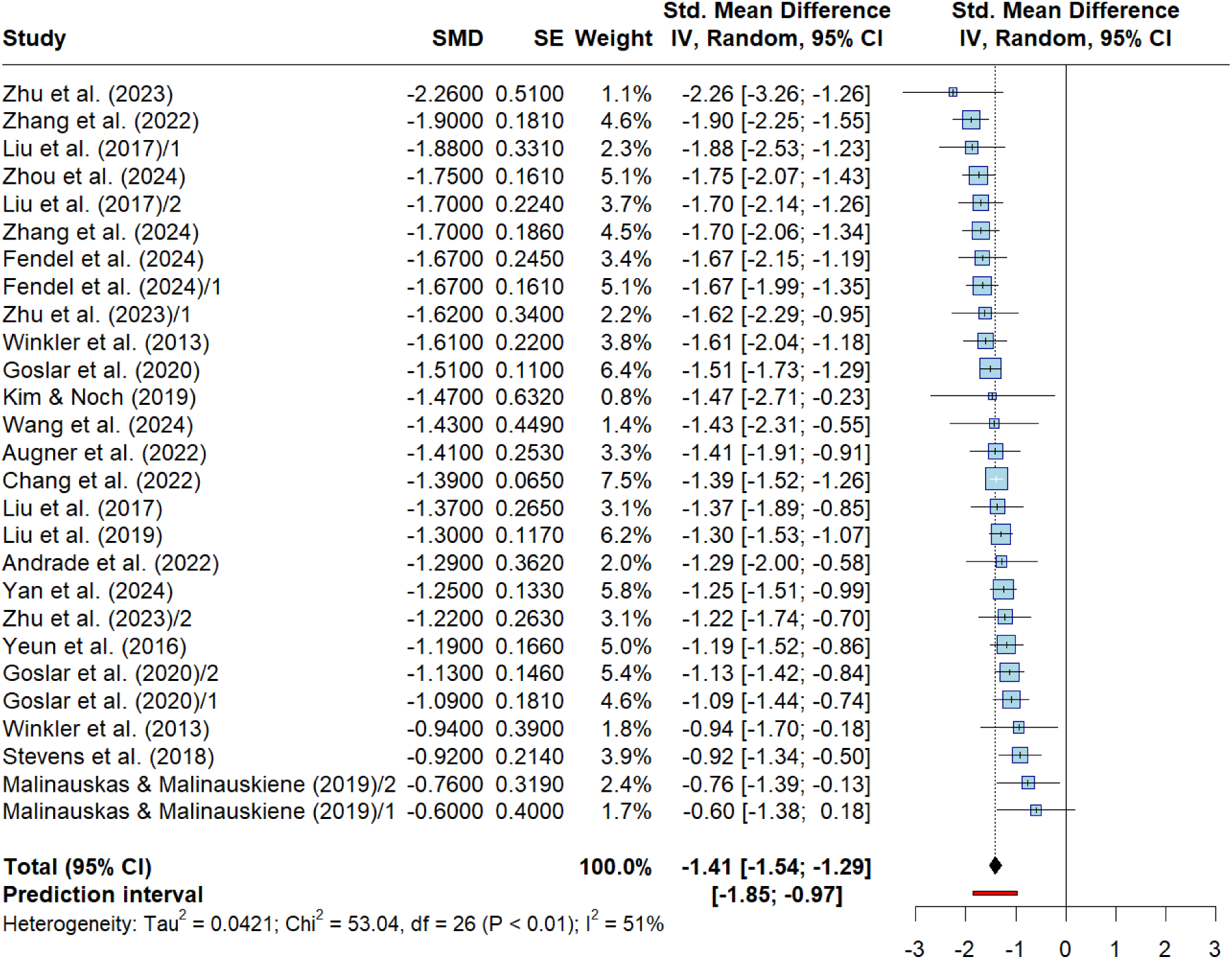
Forest plot summarizing the pooled effect sizes from meta-analyses on PUI treatments. This plot presents a synthesis of effect sizes across multiple meta-analyses after removing outliers. It visualizes the overall estimated impact of interventions for problematic internet use, while accounting for study variability and heterogeneity through a random-effects model.

**Figure 4:**
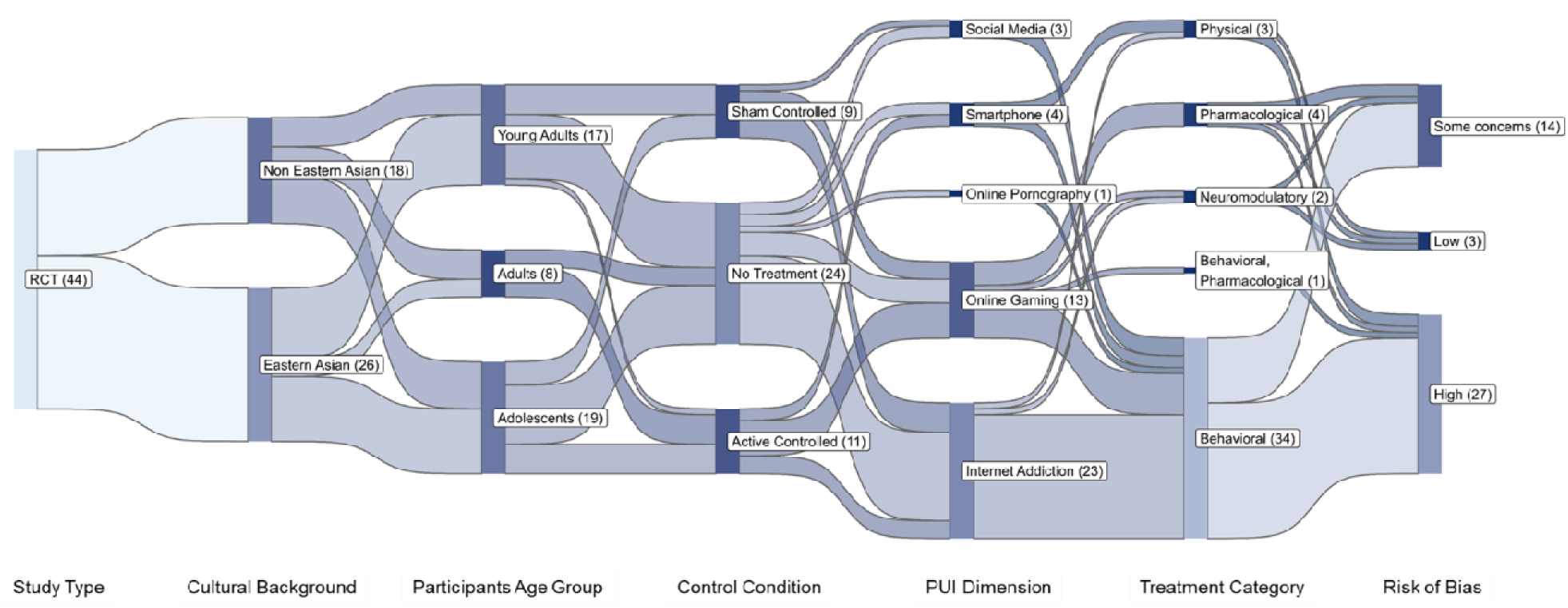
Sankey plot illustrating the characteristics of randomized controlled trials (RCTs) included in the analysis. This figure visualizes the flow of study features across multiple dimensions, including study type, cultural background, participants’ age group, control condition, PUI dimension, treatment category, and risk of bias. The gradient color intensity reflects the relative frequency of studies within each node, with darker tones indicating higher counts.

### Quality Assessment and Risk of Bias for Meta-Analyses

The methodological quality of the included meta-analyses was evaluated using the AMSTAR 2.0 checklist. Most studies were rated as low quality, with a smaller proportion classified as critically low. These results highlight notable variability in methodological rigor and underscore the need for more standardized and robust practices in future evidence syntheses. Detailed results of the AMSTAR 2.0 assessment are presented in **Error! Reference source not found.**.

In addition, we conducted an umbrella-level evaluation of potential publication bias as an extra safeguard, given the generally low methodological quality of the included studies. Funnel plot inspection (**Error! Reference source not found.**) and Egger’s regression test indicated no significant evidence of asymmetry (t = –0.09, df = 27, p = 0.93), with a bias estimate of –0.05 (SE = 0.57). These results suggest that publication bias was not a major concern in our analysis, lending further confidence to the robustness of the synthesized evidence.

### Subgroup Analysis and Meta-Regression for Meta-Analyses

Due to the overlap in PUI dimensions and treatment categories across the included meta-analyses, subgroup analysis was limited to participant groups: mixed populations (n = 20), adolescents (n = 3), young adults (n = 3), and children (n = 1). The pooled effect sizes across these subgroups were consistent, ranging from -1.45 (95% CI: [-1.59; -1.31]) for mixed groups to -1.1 (95% CI: [-2.7; 0.45]) for adolescents. Heterogeneity estimates (I²) were calculated as an additional safeguard to assess the consistency of findings across participant groups, given the variability in methodological rigor among the included meta-analyses. I² was moderate for the mixed group (43.5%) and higher for adolescents (84.2%) and young adults (53.3%), while heterogeneity was not quantifiable for the children subgroup due to a single-study group. The test for subgroup differences revealed no statistically significant differences between groups (Q = 2.79, p = 0.43), suggesting that participant type did not meaningfully contribute to variability in effect sizes (See **Error! Reference source not found.** for more details).

A significant negative association was found between the publishing year and the SMD (F(df1 = 1, df2 = 25) = 4.09, p = 0.04), suggesting that more recent meta-analyses report smaller effect sizes (**Error! Reference source not found.**-left). For the number of participants, the meta-regression showed that studies with more participants tend to report smaller effect sizes (F(df1 = 1, df2 = 25) = 6.50, p = 0.017) (**Error! Reference source not found.**-middle). However, the journal Impact Factor (IF) was not related to the SMD (F(df1 = 1, df2 = 25) = 0.16, p = 0.67) (**Error! Reference source not found.**-right).

### Meta-Analysis of RCTs (Phase 2)

#### Characteristics of Included RCTs

The initial search identified 756 studies from the included meta-analyses. After removing duplicates (n = 370), 386 unique studies were available for review. A total of 177 studies were not retrieved due to the absence of an abstract, leaving 209 studies to be considered further. Following an additional screening process, studies were excluded for the following reasons: not focused on PUI (n = 7), not addressing online aspects (n = 17), not involving a treatment (n = 11), non-English language (n = 41), not peer-reviewed (n = 17), and not clinical trials (n = 16). A total of 109 studies were excluded, resulting in 100 studies deemed eligible. Of these, 58 RCTs met the criteria for inclusion in the meta-analysis. However, eight RCTs were excluded due to insufficient data, and six RCTs were excluded because their experimental and control groups did not both consist of participants with PUI, i.e., they did not compare patient vs. patient groups. Thus, 44 RCTs were included in the final meta-analysis, providing data for 115 units of analysis. Details of the trial screening results can be found on the project’s OSF page at https://osf.io/8uc2w/.

The included RCTs included a total of 5332 participants, were conducted from 2008 to 2024, and reflected an increasing focus on PUI research in recent years. A notable rise in studies occurred from 2016 onward, with the highest numbers published in 2022 and 2023 (7 studies each). Geographically, most studies were conducted in East Asia, with China contributing 17 and Korea nine studies, followed by Iran, Turkey, and Germany (5 each). Other countries contributed smaller numbers, underscoring the global attention to PUI, with a significant focus on Eastern Asia (26 studies, 60%). Regarding PUI dimensions, most studies (n = 23) targeted internet addiction, followed by problematic online gaming (n = 13), smartphone use (n = 4), use of social media (n = 3), and use of online pornography (n = 1) (**Error! Reference source not found.**). Age group analysis revealed that 43% of studies focused on adolescents, 39% on young adults, and 18% on adults, highlighting the higher prevalence of PUI in younger populations. Control group designs primarily employed no-treatment conditions (n = 24, 55%), with smaller proportions using active controls (n = 11, 25%) and sham controls (n = 9, 20%). Behavioral interventions dominated the field (n = 34), most often evaluated against no-treatment controls, although some incorporated active (n = 7) or sham (n = 7) comparators. Pharmacological trials (n = 6) were more evenly distributed, spanning no-treatment (n = 3), active (n = 2), and sham (n = 1) designs. Neuromodulatory studies (n = 3) relied on both no-treatment and sham controls, while a single combined behavioral–pharmacological trial adopted an active comparator. No physical interventions were reported. Overall, the predominance of no-treatment controls—especially in behavioral research—highlights the limited use of more rigorous sham-controlled designs outside pharmacological and neuromodulatory modalities. This distribution reflects an emphasis on interventions targeting affected populations, particularly adolescents and young adults in East Asian cultural contexts and across different PUI dimensions (Table 2).

**Table 2:** Overview of 44 randomized controlled trials (RCTs) included in Phase 2 of the analysis. This table presents all 58 units of analysis derived from 44 RCTs, detailing key study characteristics such as cultural background, participant groups, control conditions, and treatment categories. Additional columns describe treatment types, assessed PUI dimensions, risk of bias (measured by RoB2), and standardized mean differences (SMDs), offering a structured overview of the trials included in the secondary meta-analysis.

| Author | Units of Analysis (UoA) Description | Cultural Background | Control | Participants Group | Sample Size | PUI Dimension | Treatment Types | Risk of Bias | SMD |
| --- | --- | --- | --- | --- | --- | --- | --- | --- | --- |
| Behavioral Interventions |  |  |  |  |  |  |  |  |  |
| (Agbaria 2022) | Single UoA | Non_EA | Sham Controlled | Adolescents | 160 | Internet Addiction | Cognitive Behavioral Therapy (CBT) | Low | -2.1 |
| (Alavi et al. 2021) | Immediate + Follow Up | Non_EA | Sham Controlled | Young Adults | 50 | Internet Addiction | Cognitive Behavioral Therapy (CBT) | Some concerns | -2.4 |
| (Bong et al. 2021) | PUI Scale 1, 2 | EA | Active Controlled | Adolescents | 155 | Smartphone | Music Therapy | Some concerns | -0.2 |
| (Berber Çelik 2017) | Immediate + Follow Up | Non_EA | No Treatment | Adolescents | 30 | Internet Addiction | Psychoeducation | High | -1.4 |
| (Crosby and Twhig 2016) | Single UoA | Non_EA | No Treatment | Adults | 27 | Online Pornography | Acceptance and Commitment Therapy (ACT) | High | -1.7 |
| (Dieris-Hirche et al. 2023) | PUI Scale 1, 2, 3 | Non_EA | No Treatment | Adults | 180 | Internet Addiction | Cognitive Behavioral Therapy (CBT) | High | -0.6 |
| (Du et al. 2010) | Immediate + Follow Up | EA | No Treatment | Adolescents | 56 | Internet Addiction | Cognitive Behavioral Therapy (CBT) | Some concerns | -0.7 |
| (Ede et al. 2023) | Follow Up 1 + Follow Up 2 | Non_EA | No Treatment | Young Adults | 40 | Internet Addiction | Cognitive Behavioral Therapy (CBT) | High | -23.62 |
| (Gong et al. 2022) | Single UoA | EA | No Treatment | Adolescents | 21 | Internet Addiction | Physical Exercise + Counseling | High | -2.4 |
| (He et al. 2021) | PUI Scale 1, 2 | EA | Sham Controlled | Young Adults | 48 | Online Gaming | Bias modification treatment | Some concerns | -1.6 |
| (Hou et al. 2019) | PUI Scale 1, 2 | EA | No Treatment | Young Adults | 38 | Social Media | Cognitive Behavioral Therapy (CBT) | High | -1.6 |
| (Ilano et al. 2022) | Follow Up 1, 2 | Non_EA | No Treatment | Adolescents | 30 | Internet Addiction | Cognitive Behavioral Therapy (CBT) | High | -1.3 |
| (Ji and Wong 2023) | PUI Scale 1, 2, 3 + Follow Up 1, 2, 3 | EA | No Treatment | Adolescents | 68 | Online Gaming | Cognitive Behavioral Therapy (CBT) | High | -1.0 |
| (Khazaei et al. 2017) | PUI Scale 1, 2 | Non_EA | No Treatment | Young Adults | 48 | Internet Addiction | Counseling | High | -9.5 |
| (Kim 2008) | Single UoA | EA | No Treatment | Young Adults | 25 | Internet Addiction | Counseling | High | -4.8 |
| (Kochuchakalakal Kuriala and Reyes 2023) | Single UoA | EA | No Treatment | Adolescents | 40 | Online Gaming | Cognitive Behavioral Therapy (CBT) | High | -6.1 |
| (Li et al. 2017) | PUI Scale 1, 2 + immediate + Follow Up | Non_EA | Active Controlled | Adults | 30 | Online Gaming | Cognitive Behavioral Therapy (CBT) | High | -3.7 |
| (Lindenberg et al. 2022) | Follow Up 1, 2, 3 | Non_EA | No Treatment | Adolescents | 422 | Online Gaming | Cognitive Behavioral Therapy (CBT) | Some concerns | 0.0 |
| (Liu et al. 2021) | Single UoA | EA | Sham Controlled | Adolescents | 60 | Internet Addiction | Cognitive Behavioral Therapy (CBT) | Some concerns | -0.9 |
| (Lu et al. 2020) | Treatment 1 | EA | Active Controlled | Young Adults | 95 | Smartphone | Physical Exercise | High | -8.5 |
| (Lu et al. 2020) | Treatment 2 | EA | Active Controlled | Young Adults | 95 | Smartphone | Cognitive Behavioral Therapy (CBT) | High | -8.7 |
| (Manwong et al. 2018) | PUI Scale 1, 2, 3 + Immediate + Follow Up | EA | Sham Controlled | Adolescents | 245 | Social Media | Cognitive Behavioral Therapy (CBT) | High | -0.1 |

Dual-Level Meta-Analysis of PUI Treatments
| Author | Units of Analysis (UoA) Description | Cultural Background | Control | Participants Group | Sample Size | PUI Dimension | Treatment Types | Risk of Bias | SMD |
| --- | --- | --- | --- | --- | --- | --- | --- | --- | --- |
| (Nielsen et al. 2021) | PUI Scale 1, 2 + Follow Up 1, 2 | Non_EA | Active Controlled | Adolescents | 42 | Online Gaming | Multidimensional Family Therapy (MDFT) | Some concerns | -0.8 |
| (Park et al. 2016) | Treatment 1 | EA | Active Controlled | Adolescents | 86 | Online Gaming | Cognitive Behavioral Therapy (CBT) | Some concerns | -1.3 |
| (Peng et al. 2021) | Treatment 2 | EA | Active Controlled | Adults | 112 | Internet Addiction | Cognitive Behavioral Therapy (CBT) | Low | -2.7 |
| (Shadbad 2017) | Single UoA | Non_EA | No Treatment | Adolescents | 24 | Internet Addiction | Mindfulness-based cognitive group therapy | High | -6.3 |
| (Siste et al. 2022) | PUI Scale 1, 2 | Non_EA | Active Controlled | Adults | 40 | Internet Addiction | Dialectical Behavioral Therapy | Some concerns | 0.3 |
| (Song et al. 2016) | Treatment 1 | EA | No Treatment | Young Adults | 119 | Online Gaming | Counseling | High | -1.5 |
| (Taş and Ayas 2018) | Immediate + Follow Up | Non_EA | No Treatment | Adolescents | 24 | Internet Addiction | Psychoeducation | High | -2.1 |
| (Throuvala et al. 2020) | PUI Scale 1, 2 | Non_EA | No Treatment | Young Adults | 143 | Smartphone | Brief mindfulness, Self-monitoring, and Self-exclusion | Some concerns | -0.4 |
| (Torres-Rodríguez et al. 2018) | Immediate | Non_EA | Active Controlled | Adolescents | 31 | Online Gaming | Individualized psychotherapy treatment for IGD (PIPATIC program) | Some concerns | -2.2 |
| (Wölfling et al. 2019) | PUI Scale 1 + Immediate | Non_EA | No Treatment | Adults | 143 | Internet Addiction | Cognitive Behavioral Therapy (CBT) | High | -1.4 |
| (Yaman and Yılmaz 2024) | Single UoA | Non_EA | No Treatment | Young Adults | 105 | Social Media | Psychoeducation | Some concerns | -0.5 |
| (Yang et al. 2022) | PUI Scale 1, 2 + immediate + Follow Up 1, 2 | EA | Sham Controlled | Young Adults | 44 | Internet Addiction | Cognitive Behavioral Therapy (CBT) | High | -0.4 |
| (Zeidi et al. 2020) | PUI Scale 1, 2 | Non_EA | No Treatment | Young Adults | 80 | Internet Addiction | Cognitive Behavioral Therapy (CBT) | High | -3.1 |
| (Zhang et al. 2020) | Immediate + Follow Up | EA | No Treatment | Young Adults | 18 | Internet Addiction | Counseling | High | -1.7 |
| (Zhao and Pan 2022) | PUI Scale 1, 2 | EA | Sham Controlled | Adolescents | 100 | Internet Addiction | Counseling | High | -2.7 |
| (Zheng et al. 2022) | Treatment 1 + PUI Scale 1, 3 + Immediate + Follow Up | EA | No Treatment | Adolescents | 80 | Internet Addiction | Cognitive Behavioral Therapy (CBT) | High | -0.1 |
| (Zheng et al. 2022) | Treatment 2 + PUI Scale 1, 3 + Immediate + Follow Up | EA | No Treatment | Adolescents | 80 | Internet Addiction | Cognitive Behavioral Therapy (CBT) | High | -0.2 |
| (Zheng et al. 2022) | Treatment 3 + PUI Scale 1, 3 + Immediate + Follow Up | EA | No Treatment | Adolescents | 80 | Internet Addiction | Cognitive Behavioral Therapy (CBT) | High | -0.6 |
| (Zhu et al. 2012) | Treatment 1 | EA | Active Controlled | Adults | 112 | Internet Addiction | Psychoeducation | High | -2.7 |
| Pharmacological Interventions |  |  |  |  |  |  |  |  |  |
| (Han and Renshaw 2012) | PUI Scale 1, 2 + Immediate + Follow Up | EA | Sham Controlled | Young Adults | 50 | Online Gaming | Antidepressants (Bupropion) | Some concerns | -1.0 |
| (Nam et al. 2017) | Treatment 1 | EA | Active Controlled | Adults | 30 | Online Gaming | Antidepressants (Bupropion) | Some concerns | -1.6 |
| (Nam et al. 2017) | Treatment 2 | EA | Active Controlled | Adults | 30 | Online Gaming | Antidepressants (Escitalopram) | Some concerns | -1.4 |
| (Park et al. 2016) | Treatment 2 | EA | Active Controlled | Adolescents | 86 | Online Gaming | Antidepressants (Atomoxetine) | Some concerns | -1.1 |
| (Song et al. 2016) | Treatment 2 | EA | No Treatment | Young Adults | 119 | Online Gaming | Antidepressants (Sertraline) | High | -1.2 |
| Neuromodulatory Interventions |  |  |  |  |  |  |  |  |  |
| (Jeong et al. 2020) | PUI Scale 1, 2 | EA | Sham Controlled | Young Adults | 26 | Online Gaming | tDCS | High | -0.1 |
| (Peng et al. 2021) | Treatment 1 | EA | Active Controlled | Adults | 112 | Internet Addiction | Electroacupuncture | Low | -3.1 |
| (Peng et al. 2021) | Treatment 3 | EA | Active Controlled | Adults | 112 | Internet Addiction | Electroacupuncture | Low | -4.5 |

Dual-Level Meta-Analysis of PUI Treatments
| Author | Units of Analysis (UoA) Description | Cultural Background | Control | Participants Group | Sample Size | PUI Dimension | Treatment Types | Risk of Bias | SMD |
| --- | --- | --- | --- | --- | --- | --- | --- | --- | --- |
| (Zhu et al. 2012) | Treatment 2 | EA | Active Controlled | Adults | 112 | Internet Addiction | Electroacupuncture | High | -3.1 |
| **Physical Interventions** |  |  |  |  |  |  |  |  |  |
| (Xiao et al. 2021) | Treatment 1 + Immediate + Follow Up | EA | No Treatment | Young Adults | 96 | Smartphone | Group-based baduanjin exercise | High | -1.1 |
| (Xiao et al. 2021) | Treatment 2 + Immediate + Follow Up | EA | No Treatment | Young Adults | 96 | Smartphone | Group-based basketball exercise | High | -1.4 |
| (Zhang et al. 2023) | Treatment 1 | EA | No Treatment | Young Adults | 93 | Internet Addiction | General Exercise | Low | -0.9 |
| (Zhang et al. 2023) | Treatment 2 | EA | No Treatment | Young Adults | 93 | Internet Addiction | Taichi Exercise | Low | -0.4 |
| (Zhang et al. 2024) | Treatment 1 | EA | No Treatment | Young Adults | 90 | Smartphone | General Exercise | Low | -0.8 |
| (Zhang et al. 2024) | Treatment 2 | EA | No Treatment | Young Adults | 90 | Smartphone | General Exercise | Low | -0.5 |
| **Combined Interventions** |  |  |  |  |  |  |  |  |  |
| (Kim et al. 2012) | PUI Scale 1, 2 + immediate + Follow Up | EA | Active Controlled | Adolescents | 65 | Online Gaming | Cognitive Behavioral Therapy (CBT) + Antidepressants (Bupropion) | High | -0.8 |
| (Zhu et al. 2012) | Treatment 3 | EA | Active Controlled | Adults | 112 | Internet Addiction | Psychoeducation + Electroacupuncture | High | -4.5 |

The analysis of the frequency with which each RCT appeared across the 20 meta-analyses reveal minimal overlap among the included studies. The most frequently cited study (Du et al. 2010) was included in only eight of the meta-analyses, while many other studies were represented in just one or two. The 115 units of analysis identified across the 44 RCTs were categorized into various PUI scale categories, with a predominant focus on symptom scales (91 units, 79%), followed by consumption (14 units, 12%) and craving (10 units, 9%) scales. In terms of treatment categories, behavioral treatments were most commonly used (82 units, 76%), followed by pharmacological (10 units, 9%), physical (8 units, 7%), and neuromodulatory (4 units, 3%) treatments. Combined treatment approaches included four units of behavioral and pharmacological treatments and two units of behavioral and neuromodulatory treatments. This distribution reflects a significant emphasis on behavioral interventions for PUI, with some inclusion of other treatment types.

### Analytic Results of RCTs

As this analysis is focused on the impact of different treatment approaches to PUI, we first combined units of analysis over treatment approaches. This means that units of analysis that represent different PUI assessment scales and/or different measurement times (i.e., immediate or follow-ups) were combined. This resulted in 56 units of analysis, on which we conducted a random-effects meta-analysis. The pooled SMD was -1.96 (95% CI [-2.39; -1.33], t= -6.55, p < 0.0001), reflecting a moderate to large effect size favoring treatment. However, substantial heterogeneity was observed (I² = 95.0% [94.1%; 95.7%]), with a significant test for heterogeneity (Q = 1129.36, p < 0.0001), suggesting considerable variability across the studies.

To further explore the sources of this heterogeneity, we conducted subgroup analyses based on PUI dimensions and treatment categories. These analyses did not show a significant change in heterogeneity among different PUI dimensions (**Error! Reference source not found.**a). For different treatment approaches, a reduction was observed only for pharmacological treatments (I^2^ = 0% among seven units of analysis out of four studies, p = 0.61) and physical exercise treatments (I^2^ = 51% among six units of analysis out of three studies, p = 0.07) (**Error! Reference source not found.**b). Using a combination of PUI dimensions and treatment approaches also showed some reduction in heterogeneity, but only among a smaller group of units of analysis (**Error! Reference source not found.**). Thus, despite some reductions in some subgroups, high I² values within other subgroups indicated persistent heterogeneity and suggest that outlier analysis is necessary to investigate whether extreme values are contributing to the observed variability.

We have the same outlier removal process as used in the first phase, in addition to a single study (Khazaei et al. 2017), as the reported effect size was out of the range. This process found 32 outliers, leaving 26 units of analysis for further steps. This led to a drop in the pooled effect size (SDD = -1.77 (95% CI: [-2.03; -1.50], t=-13.73, p<0.0001)), and a reduction in the heterogeneity index (I^2^ = 73.5% [61.0%; 82.0%]) (Figure 5). We then conducted a subgroup analysis considering the combination of PUI dimensions and treatment approaches to explore sources of heterogeneity. The test for subgroup differences was significant (Q = 19.27, df = 7, p = 0.0074), indicating that heterogeneity was influenced by how PUI dimensions and treatment categories were represented across studies. Notably, the internet addiction (behavioral) subgroup had a larger effect size with an SMD of -2.01 (95% CI: [-2.39;-1.62], p<0.01) and heterogeneity (I² = 66.0%). However, in other subgroups, although showing less heterogeneity, the effect sizes were not significant (p-value>0.05).

**Figure 5.**
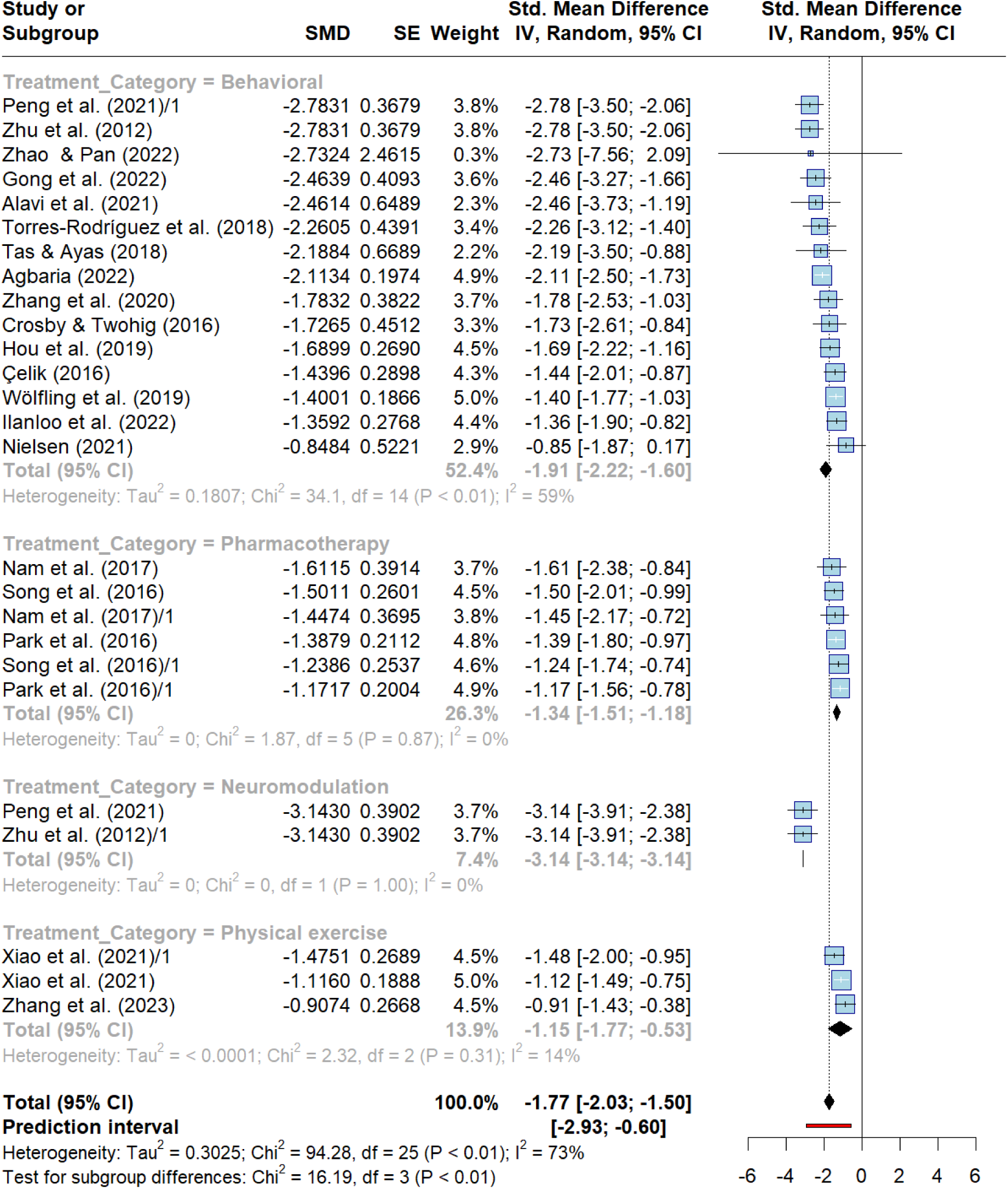
Forest plot of randomized controlled trials (RCTs), grouped by treatment category. This figure illustrates the comparative impact of different intervention types for problematic internet use, including behavioral, pharmacological, neuromodulatory, and physical exercise treatments. Studies are grouped by treatment category, and the plot reflects pooled effect sizes after outlier removal. Subgroup analyses reveal variation in impact across intervention types.

### Publication Bias and Risk of Bias Analysis for RCTs

To assess potential publication bias, we examined the funnel plot for the studies included before and after outlier removal (**Error! Reference source not found.**). Visual inspection of the funnel plot suggested some asymmetry, which was further quantified using Egger’s regression test. The test revealed significant funnel plot asymmetry before removal of outliers (t = -7.74, df = 56, p-value < 0.0001) and still some evidence but weaker (t = -2.54, df = 24, p-value = 0.0178) after removal of outliers, indicating the presence of small-study effects or publication bias at a moderate level (bias estimate= -2.4060, SE = 0.9460).

The risk of bias was evaluated using the RoB2 tool (Sterne et al. 2019), across five domains: (D1) bias arising from the randomization process, (D2) bias due to deviations from intended interventions, (D3) bias due to missing outcome data, (D4) bias in the measurement of the outcome, and (D5) bias in the selection of the reported result. Studies were categorized as having “low risk,” “some concerns,” or “high risk” based on these dimensions, with an overall risk assigned accordingly. Of the 44 RCTs included, only four studies (9%) were rated as having an overall low risk of bias, while 14 studies (32%) had some concerns, and 28 studies (59%) were rated as having a high risk of bias. The most frequent sources of bias were in D4 (bias in outcome measurement) and D5 (bias in the selection of reported results), with a significant proportion of studies showing “some concerns” or “high risk” in these dimensions. Conversely, D3 (bias due to missing outcome data) exhibited the lowest levels of bias, with most studies rated as low risk in this domain.

A detailed breakdown of risk of bias assessments by study and dimension is provided in **Error! Reference source not found..**

### Subgroup Analysis and Meta-Regression in RCTs

We restricted our subgroup and meta-regression analysis to the 26 units of analysis that remained after outlier removal. In this regard, despite some small differences, cultural background (non-Eastern vs. Eastern), PUI scale category (symptoms vs. consumption), and risk of bias (low, some concerns, and high) did not show significant differences in effect sizes (p-values: 0.59, 0.92, and 0.20, respectively). On the other hand, control condition significantly affected the effect size (p-value < 0.0001) where sham-controlled studies (n=3) showed higher effect (SMD = -2.14, 95% CI: [-2.47; -1.8221]) compared to no treatment (n=13, SMD = -1.43, 95% CI: [-1.6373; -1.22]) and active-controlled studies (n=10, SMD= - 2.05, 95% CI: [-2.66; -1.44]). Furthermore, results showed that studies investigting adults showed stronger effects (n=8, SMD = -2.24, 95% CI: [-2.89; -1.58]) compared to those involving adolescents (n=10, SMD = -1.65, 95% CI: [-2.03; -1.27]) and young adults (n=8, SMD= -1.38, 95% CI: [-1.67; - 1.08]), and this difference was significant (p-value = 0.0146). Concerning PUI dimensions, treatments of internet addiction showed the strongest effects (n=14, SMD=2.11, 95% CI: [-2.53; -1.68]) and problematic use of smartphones showed the lowest effects (n=2, SMD=-1.24, 95% CI: [-3.43; 0.94]), and the difference was also significant (p-value=0.0010). Finally, treatment categories were also significant (p-value = 0.0010), with neuromodulatory treatments having the highest effect (n=2, SMD = -3.14, 95% CI: [-3.14; -3.14]) and physical exercise the lowest (n=3, SMD=-1.15, 96% CI: [-1.77; -0.53]). However, caution is warranted in interpreting the findings from subgroups with very small sample sizes (e.g., n = 2 or 3), as these estimates are more susceptible to random error and may reflect exaggerated effects due to limited statistical power. See **Error! Reference source not found.** for the details.

Finally, we investigated the role of publication year, sample size, frequency of appearance in meta-analyses, number of days of treatment, and number of sessions of treatment. Results showed that publication year and sample size did not significantly predict effect sizes (p-values = 0.2814 and 0.5433, respectively). However, the frequency of appearance in meta-analyses significantly predicted effect sizes (F(1, k–2) = 4.63, p = 0.0418), accounting for 20.58% of the between-study variance. This means that the more an RCT was included in meta-analyses, the lower the effect. Additionally, while the number of days did not predict effect sizes (p-value = 0.2072), the number of sessions showed marginally significant prediction power (F(1, k–2) = 3.6557, p = 0.0719), accounting for 14.07% of the between-study variance. See **Error! Reference source not found.** for more details.

The meta-regression analyses examined the effect of several moderators on effect sizes. Publication year did not show a significant association (b=0.03 (p = 0.416)). Similarly, sample size had no significant effect (b=−0.0024, p = 0.416). However, frequency, defined as the number of meta-analyses identified in the study, was found to have a significant negative relationship with effect sizes, b=−0.31 (p = 0.046), indicating that studies identifying a higher number of meta-analyses tended to report smaller effect sizes. The number of treatment days (n_Days) and the number of treatment sessions (n_Sessions) did not significantly explain the variability in effect sizes, of b=0.0030 (p = 0.3515) and b=0.0194 (p = 0.1505), respectively. Detailed results are provided in **Error! Reference source not found.**.

## 4 Discussion

This study provides a comprehensive evaluation of treatments for PUI by analyzing evidence from two levels: (1) meta-analyses (phase 1) and (2) RCTs (phase 2). This dual-level analysis allows for a deeper understanding of the state of research, highlighting consistencies and discrepancies between aggregated evidence (meta-analyses) and primary-level data (RCTs).

### Scope of Studies and Dimensions of PUI

The analysis of 20 meta-analyses (phase 1) suggests that meta-analyses primarily focused on internet addiction, problematic smartphone use, problematic online gaming, and problematic social media usage, with internet addiction being the most commonly studied dimension. However, dimensions such as problematic online shopping and use of online pornography were underrepresented (Fig 2 right panel).

At the RCT level (phase 2), a similar focus on internet addiction was observed, consistent with phase 1 findings. However, individual RCTs showed slightly greater diversity, including studies on less-explored dimensions such as online shopping addiction, which are typically underrepresented in meta-analyses due to limited data. This aligns with findings from studies such as (J Kuss et al. 2014), highlighting both the growing diversity and the actual or perceived severity/prevalence of different PUI dimensions in primary research. This further support the formation of PUI as an umbrella term covering these various diagnostic constructs (Zare-Bidoky et al. 2025).

### Intervention Approaches

Both meta-analyses (phase 1) and RCTs (phase 2) highlighted the predominance of behavioral interventions, particularly those based on psychological counseling and cognitive-behavioral therapy (CBT). This alignment underscores the strong evidence base for behavioral treatments across both levels of analysis, as supported by similar findings (Xu et al. 2021).

However, a key divergence emerged in the representation of pharmacological and neuromodulatory interventions. While meta-analyses included relatively few studies testing these approaches, phase 2 revealed an increasing number of RCTs evaluating pharmacological and neuromodulatory treatments, though with mixed results. These findings align with emerging literature (de Sá, Rafael Richard Clorado et al. 2023; Xu et al. 2022), which suggests a limited but potentially promising role for pharmacological interventions in specific PUI dimensions, such as online gambling, where more primary trials are available.

### Outcome Measures and Heterogeneity

Both meta-analyses (phase 1) and RCTs (phase 2) demonstrated significant variability in outcome measures. Meta-analyses primarily relied on frequently used and psychometrically validated self-assessment scales like the Young Internet Addiction Scale (YIAS) (Young 1998) and the Smartphone Addiction Scale (SAS) (Kwon et al. 2013). Similarly, RCTs employed these scales but also introduced more exploratory measures, contributing to some heterogeneity. While the use of standardized scales facilitates comparability across studies and enables meta-analytic synthesis, the addition of novel or less-validated measures introduces variability. This highlights both the value of common measurement tools and the need for expanding validated measures to capture emerging dimensions of problematic usage of the internet (PUI).

### Observed Outcomes of Interventions

At the meta-analytic level (Phase 1), the pooled random-effects effect size (−1.41) indicated that interventions were associated with meaningful reductions in PUI. The moderately high heterogeneity, even after removing outliers (I² = 51.0%), highlighted the diversity in populations, interventions, and methodologies. Phase 2 (RCTs) similarly showed that interventions were linked to reductions in PUI (−1.77), though effect sizes were smaller and varied across study characteristics, such as age group, type of PUI, or intervention format, reflecting context-dependent outcomes. Heterogeneity remained high (I² = 73.0%).

### Risk of Bias and Methodological Concerns

The meta-analyses, assessed using AMSTAR 2.0, were predominantly of low to critically low quality, reflecting a need for more rigorous and transparent methodologies. Phase 2 (RCTs) found that many trials lacked standardized protocols, adequate blinding, or longer-term follow-up. This finding is consistent with the broader literature, including (Basenach et al. 2023), which also emphasized the importance of improving methodological quality in PUI research.

### Subgroup Analysis and Temporal Trends

Subgroup analyses in meta-analyses (phase 1) revealed no significant differences in treatment effects across mixed populations, adolescents, young adults, and children. At the RCT level (phase 2), similar findings were observed, though the smaller sample sizes in RCTs often limited subgroup analysis. The higher heterogeneity among adolescent and young adult groups was consistent across both phases, as noted in (Ayub et al. 2023; Chang et al. 2022), suggesting potential variability in intervention outcome based on age. The meta-regression findings from phase 1 showed a negative correlation between publishing year and effect size; however, this was not apparent in phase 2.

### Geographical and Cultural Contexts

The concentration of studies in Eastern Asia, observed at both the meta-analysis and RCT levels, may reflect the high prevalence of PUI in this region. Similar findings reported by other scholars (See Meng et al. 2022, for example) further support the dominant contribution of Eastern Asian research to the evidence base on PUI. However, although no significant differences in effect sizes were observed between Eastern-Asian and non-Eastern-Asian studies, expanding research to other geographical and cultural contexts would enhance the global applicability of the findings.

## 5 Conclusion

This meta-analysis synthesized findings from 20 meta-analyses and 44 RCTs, providing a comprehensive evaluation of treatments for PUI. The results revealed a moderate to large overall treatment effect, but significant heterogeneity was observed across studies, reflecting differences in intervention types, populations, outcome measures, and follow-up durations. Regarding the quality of evidence, many meta-analyses were rated as low quality, and individual studies often had methodological limitations such as inadequate blinding and small sample sizes. This combination of heterogeneity and variable quality underscores the need for more rigorous methodologies in future research, including further investigation into the specific contributions of different treatment modalities and PUI dimensions.

In consideration of the wide diversity of activities that people may have on the internet and smartphones, it seems important to consider the development and assessment of tailored interventions targeting specific PUI dimensions, such as problematic engagement in online gaming, online pornography, social media use, and online shopping. Although our analyses did not reveal consistent significant differences in efficacy across these dimensions, it is plausible that activity-specific interventions could enhance effectiveness, given that people may differ in their motivations to use such services and in the psychological characteristics contributing to PUI (Brahim et al. 2019; Zanetta Dauriat et al. 2011).

Inclusion criteria for future studies should consider new developments in the field, such as the recent inclusion of diagnostic guidelines for online gaming disorder in ICD-11 (Saunders et al. 2025) or the possible characterisation of other specified disorders due to addictive behaviors (Brand et al. 2025; Brand et al. 2022).

Behavioral interventions—particularly Cognitive Behavioral Therapy (CBT)—have shown promising impacts. Future studies should further delineate the active components of effective interventions (e.g., cognitive restructuring, exposure, motivational approaches), identify common and specific elements by type of PUI, and assess various delivery formats (e.g., individual, group-based, face-to-face, or online). Additionally, integration with digital tools warrants exploration. Other approaches, such as pharmacological treatments and neuromodulation, should also be further investigated to clarify their indications and delivery modalities.

The quality issues observed in studies and meta-analyses on PUI can be partly attributed to the recent emergence of these phenomena, as well as the lack of consensus regarding measurement tools, diagnostic criteria, and contributing factors. The recent introduction of specific diagnostic guidelines (for online gaming, gambling, and use of pornography) by the World Health Organization, combined with advances in psychometric instruments, presents a timely opportunity to establish research-specific standards in this field.

The dynamic nature of human-digital interactions invites, within a treatment perspective, the development of more refined, adaptive, and creative assessment methods that can address the complexities of PUI at both the individual and group levels, while capturing the multidimensional nature of interactions between humans and digital devices.. Advancing such approaches will require the active involvement of individuals with lived experience, alongside careful attention to privacy and ethical considerations.

To advance the field, future research may prioritize the development of methodological checklists and consensus guidelines that enhance conceptual consistency and methodological rigor at both levels (i.e., meta-analyses and RCTs). There is a need for more studies focusing on under-explored treatment modalities, such as pharmacotherapies, and dimensions of PUI like those involving online pornography and shopping, which remain significantly under-investigated. Additionally, the role of co-occurring concerns should be more fully considered (Bullock and Potenza 2012; Potenza et al. 2019).

Overall, this meta-analysis provides evidence for efficacy of behavioral interventions, particularly CBT, and physical exercises and potentially pharmacological and neuromodulatory treatments for various aspects of PUI, while providing a roadmap for quality improvement in future trials.

## Data Availability

All data produced in the present study are available upon reasonable request to the authors

https://osf.io/8uc2w

