## Supplementry Material for "Treatment Approaches for Problematic Usage of the Internet (PUI): A Dual-Level Meta-Analysis of Meta-Analyses and Randomized Controlled Trials"

### Supplementary Material

### Search String Development

*Supplementary Table 1: Search String Development Steps*

| Category | Search Term | Results | Overlap with PUI in General |
|---|---|---|---|
| **a. PUI in General** | (“internet addiction”[tiab:~0] OR “internet usage disorder”[tiab:~0] OR “internet dependence”[tiab:~0] OR “problematic internet”[tiab:~0] OR “pathological internet”[tiab:~0] OR “online addiction”[tiab:~0] OR “compulsive internet” [tiab:~0] OR “internet overuse”[tiab:~0] OR “Internet disorder”[tiab:~0] OROR ”smartphone addiction”[tiab:~0] OR “phone addiction”[tiab:~0]) | **3,677** | **3,677** |
| **b. Online Gaming** | ((patholog*[Title/Abstract] OR problem*[Title/Abstract] OR addict*[Title/Abstract] OR compulsive[Title/Abstract] OR dependen*[Title/Abstract] OR disorder*[Title/Abstract])<br><br>AND<br><br>(video[Title/Abstract] OR computer[Title/Abstract])<br><br>AND<br><br>(gaming[Title/Abstract] OR game[Title/Abstract])<br><br>AND<br><br>(internet[Title/Abstract] OR online[Title/Abstract] OR cyber*[Title/Abstract] OR digital[Title/Abstract] OR virtual[Title/Abstract])) | **822** | **153** |
| **c. Online Gambling** | ((internet[Title/Abstract] OR online[Title/Abstract] OR cyber*[Title/Abstract])<br><br>AND<br><br>(gambl*[Title/Abstract] OR gambl*[MeSH])) | **1478** | **264** |
| **d. Online Buying-Shopping Disorder** | ((internet[Title/Abstract] OR online[Title/Abstract] OR cyber*[Title/Abstract] OR electronic[Title/Abstract] OR digital[Title/Abstract])<br><br>AND | **192** | **64** |

| Category | Search Term | Results | Overlap with PUI in General |
|---|---|---|---|
| | (“buying addiction”[tiab:~0] OR “addictive buying”[tiab:~0] OR “compulsive buying”[tiab:~0] OR “impulsive buying”[tiab:~0] OR “problematic buying”[tiab:~0] OR “pathological buying”[tiab:~0] OR “excessive buying”[tiab:~0] OR “compensatory buying”[tiab:~0] OR “obsessive buying”[tiab:~0] OR “buying disorder”[tiab:~0] OR “shopping addiction”[tiab:~0] OR “addictive shopping”[tiab:~0] OR “compulsive shopping”[tiab:~0] OR “impulsive shopping”[tiab:~0] OR “problematic shopping”[tiab:~0] OR “pathological shopping”[tiab:~0] OR “excessive shopping”[tiab:~0] OR “compensatory shopping”[tiab:~0] OR “obsessive shopping”[tiab:~0] OR “shopping disorder”[tiab:~0] OR “spending addiction”[tiab:~0] OR “addictive spending”[tiab:~0] OR “compulsive spending”[tiab:~0] OR “impulsive spending”[tiab:~0] OR “problematic spending”[tiab:~0] OR “pathological spending”[tiab:~0] OR “excessive spending”[tiab:~0] OR “compensatory spending”[tiab:~0] OR “obsessive spending”[tiab:~0] OR “spending disorder”[tiab:~0] OR “purchasing addiction”[tiab:~0] OR “addictive purchase”[tiab:~0] OR “compulsive purchase”[tiab:~0] OR “impulsive purchase”[tiab:~0] OR “problematic purchase”[tiab:~0] OR “pathological purchase”[tiab:~0] OR “excessive purchase”[tiab:~0] OR “compensatory purchase”[tiab:~0] OR “obsessive purchase”[tiab:~0] OR “purchasing disorder”[tiab:~0] OR “buying problem”[tiab:~0] OR “shopping problem”[tiab:~0] OR “spending problem”[tiab:~0] OR “purchasing problem”[tiab:~0] OR shopaholic[Title/Abstract] OR oniomania[Title/Abstract] OR overshoping[Title/Abstract] OR overspending[Title/Abstract]) ) | | |
| **e. Cyberchondria** | (cyberchondria[Title/Abstract] OR cyberchondriasis[Title/Abstract]<br><br>OR<br><br>((online[Title/Abstract] OR internet[Title/Abstract] OR web[Title/Abstract])<br><br>AND<br><br>(hypochondriasis[Title/Abstract] OR hypochondria[Title/Abstract])) | **218** | **31** |
| **f. Online Compulsive Sexual Behavior Disorder** | ( “online sex”[tiab:~0] OR sexting[Title/Abstract] OR “online sex disorder”[tiab:~0] OR porn*[Title/Abstract] OR Cyberporn*[Title/Abstract] OR Cybersex*[Title/Abstract]) | **2,523** | **138** |
| **g. Cyberbullying** | ((Internet[Title/Abstract] OR Online[Title/Abstract] OR Cyber*[Title/Abstract])<br><br>AND | **1,434** | **60** |

| Category | Search Term | Results | Overlap with PUI in General |
|---|---|---|---|
| | bully*[Title/Abstract])) | | |
| **f. Problematic Social Media Use** | (“Social media addiction”[tiab:~0] OR “Social media problematic use”[tiab:~0] OR “problematic Social media use”[tiab:~0] OR “Social media disorder”[tiab:~0] OR “Social media abuse”[tiab:~0] OR "problematic usage of the internet"[tiab:~0] OR "problematic usage of internet"[tiab:~0] OR “Social media misuse”[tiab:~0] OR “Social media compulsive use”[tiab:~0] OR ”Compulsive Use of Social Media”[tiab:~0] OR ”Excessive Social Media use”[tiab:~0] OR “social media addiction”[tiab:~0] OR “social networking site dependence”[tiab:~0] OR “social media dependence”[tiab:~0] OR “pathological social networking site use”[tiab:~0] OR “pathological social media use”[tiab:~0] OR “compulsive social networking site use”[tiab:~0] OR “compulsive social media use”[tiab:~0] OR “excessive social networking site use”[tiab:~0] OR “excessive social media use”[tiab:~0]) | **548** | **130** |
| **g. Digital Hoarding** | ((Internet[Title/Abstract] OR Online[Title/Abstract] OR Cyber*[Title/Abstract] OR Electronic[Title/Abstract] OR Digital[Title/Abstract] OR Virtual[Title/Abstract])<br><br>AND<br><br>hoard*[Title/Abstract])) | **138** | **8** |
| **h. Intervention** | (Medication[Title/Abstract] OR treat*[Title/Abstract] OR Interven*[Title/Abstract] OR Therap*[Title/Abstract] OR Pharmacotherapy[Title/Abstract] OR Pharmacological[Title/Abstract] OR psychotherap*[Title/Abstract] OR training[Title/Abstract] OR Curricul*[Title/Abstract] OR Counsel*[Title/Abstract] OR Educ*[Title/Abstract] OR Psychoeduc*[Title/Abstract] OR Workshop*[Title/Abstract] OR Non-Pharmacological[Title/Abstract] OR psychopharma*[Title/Abstract] OR “Cognitive Behavioral Therapy”[tiab:~0] OR CBT[Title/Abstract] OR “Cognitive Therapy”[tiab:~0] OR “Behavioral Therapy”[tiab:~0] OR Exercise[Title/Abstract] OR “Physical Activity”[tiab:~0] OR “12 step”[tiab:~0] OR “Twelve step”[tiab:~0] OR Self-help[Title/Abstract] OR Anonymous[Title/Abstract] OR Psychoanalytic[Title/Abstract] OR Psychodynamic[Title/Abstract] OR Neuromodulation[Title/Abstract] OR Neurostimulation[Title/Abstract] OR tDCS[Title/Abstract] OR “transcranial direct current stimulation”[tiab:~0] OR tACS[Title/Abstract] OR “transcranial alternating current stimulation”[tiab:~0] OR tRNS[Title/Abstract] OR “transcranial random noise stimulation”[tiab:~0] OR TMS[Title/Abstract] OR “transcranial magnetic stimulation”[tiab:~0] OR TBS[Title/Abstract] OR “theta burst stimulation”[tiab:~0] OR Manual[Title/Abstract] OR “Case Series”[tiab:~0] OR Randomized[Title/Abstract] OR Trial[Title/Abstract] OR Controlled[Title/Abstract]) | **11,415,861** | **NA** |

| Category | Search Term | Results | Overlap with PUI in General |
|---|---|---|---|
| **Combined Terms** | (a OR b OR c OR d OR e OR f OR g) AND h | **4,760** | |

The above mentioned search strategy resulted in the search criteria as follows: (("Internet addiction"[tiab:~0] OR "internet usage disorder"[tiab:~0] OR "internet dependence"[tiab:~0] OR "problematic internet"[tiab:~0] OR "pathological internet"[tiab:~0] OR "online addiction"[tiab:~0] OR "compulsive internet" [tiab:~0] OR "internet overuse"[tiab:~0] OR "Internet disorder"[tiab:~0] OR "smartphone addiction"[tiab:~0] OR "phone addiction"[tiab:~0])) OR ((patholog*[Title/Abstract] OR problem*[Title/Abstract] OR addict*[Title/Abstract] OR compulsive[Title/Abstract] OR dependen*[Title/Abstract] OR disorder*[Title/Abstract]) AND (video[Title/Abstract] OR computer[Title/Abstract]) AND (gaming[Title/Abstract] OR game[Title/Abstract]) AND (Internet[Title/Abstract] OR Online[Title/Abstract] OR Cyber*[Title/Abstract] OR Digital[Title/Abstract] OR Virtual[Title/Abstract])) OR ((Internet[Title/Abstract] OR Online[Title/Abstract] OR Cyber*[Title/Abstract]) AND (gambl*[Title/Abstract] OR gambl*[MeSH])) OR ((Internet[Title/Abstract] OR Online[Title/Abstract] OR Cyber*[Title/Abstract] OR Electronic[Title/Abstract] OR Digital[Title/Abstract]) AND ("buying addiction"[tiab:~0] OR "addictive buying"[tiab:~0] OR "compulsive buying"[tiab:~0] OR "impulsive buying"[tiab:~0] OR "problematic buying"[tiab:~0] OR "pathological buying"[tiab:~0] OR "excessive buying"[tiab:~0] OR "compensatory buying"[tiab:~0] OR "obsessive buying"[tiab:~0] OR "buying disorder"[tiab:~0] OR "shopping addiction"[tiab:~0] OR "addictive shopping"[tiab:~0] OR "compulsive shopping"[tiab:~0] OR "impulsive shopping"[tiab:~0] OR "problematic shopping"[tiab:~0] OR "pathological shopping"[tiab:~0] OR "excessive shopping"[tiab:~0] OR "compensatory shopping"[tiab:~0] OR "obsessive shopping"[tiab:~0] OR "shopping disorder"[tiab:~0] OR "spending addiction"[tiab:~0] OR "addictive spending"[tiab:~0] OR "compulsive spending"[tiab:~0] OR "impulsive spending"[tiab:~0] OR "problematic spending"[tiab:~0] OR "pathological spending"[tiab:~0] OR "excessive spending"[tiab:~0] OR "compensatory spending"[tiab:~0] OR "obsessive spending"[tiab:~0] OR "spending disorder"[tiab:~0] OR "purchasing addiction"[tiab:~0] OR "addictive purchase"[tiab:~0] OR "compulsive purchase"[tiab:~0] OR "impulsive purchase"[tiab:~0] OR "problematic purchase"[tiab:~0] OR "pathological purchase"[tiab:~0] OR "excessive purchase"[tiab:~0] OR "compensatory purchase"[tiab:~0] OR "obsessive purchase"[tiab:~0] OR "purchasing disorder"[tiab:~0] OR "buying problem"[tiab:~0] OR "shopping problem"[tiab:~0] OR "spending problem"[tiab:~0] OR "purchasing problem"[tiab:~0] OR shopaholic[Title/Abstract] OR oniomania[Title/Abstract] OR overshoping[Title/Abstract] OR overspending[Title/Abstract]) ) OR ( cyberchondria[Title/Abstract] OR cyberchondriasis[Title/Abstract] OR ( (online[Title/Abstract] OR internet[Title/Abstract] OR web[Title/Abstract]) AND (hypochondriasis[Title/Abstract] OR hypochondria[Title/Abstract]) ) OR ( "online sex"[tiab:~0] OR sexting[Title/Abstract] OR "online sex disorder"[tiab:~0] OR porn*[Title/Abstract] OR Cyberporn*[Title/Abstract] OR Cybersex*[Title/Abstract]) OR ((Internet[Title/Abstract] OR Online[Title/Abstract] OR Cyber*[Title/Abstract]) AND bully*[Title/Abstract]) OR ( "Social media addiction"[tiab:~0] OR "Social media problematic use"[tiab:~0] OR "problematic Social media use"[tiab:~0] OR "Social media disorder"[tiab:~0] OR "Social media abuse"[tiab:~0] OR "problematic usage of the internet"[tiab:~0] OR "problematic usage of internet"[tiab:~0] OR "Social media misuse"[tiab:~0] OR "Social media compulsive use"[tiab:~0] OR "Compulsive Use of Social Media"[tiab:~0] OR "Excessive Social Media use"[tiab:~0] OR "social media addiction"[tiab:~0] OR "social networking site dependence"[tiab:~0] OR "social media dependence"[tiab:~0] OR "pathological social networking site use"[tiab:~0] OR "pathological social media use"[tiab:~0] OR "compulsive social networking site use"[tiab:~0] OR "compulsive social media use"[tiab:~0] OR "excessive social networking site use"[tiab:~0] OR "excessive social media use"[tiab:~0]) OR ( (Internet[Title/Abstract] OR Online[Title/Abstract] OR Cyber*[Title/Abstract] OR Electronic[Title/Abstract] OR Digital[Title/Abstract] OR Virtual[Title/Abstract]) AND hoard*[Title/Abstract]) ) AND (Medication[Title/Abstract] OR treat*[Title/Abstract] OR Interven*[Title/Abstract] OR Therap*[Title/Abstract] OR Pharmacotherapy[Title/Abstract] OR Pharmacological[Title/Abstract] OR psychotherap*[Title/Abstract] OR training[Title/Abstract] OR Curricul*[Title/Abstract] OR Counsel*[Title/Abstract] OR Educ*[Title/Abstract] OR Psychoeduc*[Title/Abstract] OR Workshop*[Title/Abstract] OR Non-Pharmacological[Title/Abstract] OR psychopharma*[Title/Abstract] OR "Cognitive Behavioral Therapy"[tiab:~0] OR CBT[Title/Abstract] OR "Cognitive Therapy"[tiab:~0] OR "Behavioral Therapy"[tiab:~0] OR Exercise[Title/Abstract] OR

"Physical Activity"[tiab:~0] OR "12 step"[tiab:~0] OR "Twelve step"[tiab:~0] OR Self-help[Title/Abstract] OR Anonymous[Title/Abstract] OR Psychoanalytic[Title/Abstract] OR Psychodynamic[Title/Abstract] OR Neuromodulation[Title/Abstract] OR Neurostimulation[Title/Abstract] OR tDCS[Title/Abstract] OR "transcranial direct current stimulation"[tiab:~0] OR tACS[Title/Abstract] OR "transcranial alternating current stimulation"[tiab:~0] OR tRNS[Title/Abstract] OR "transcranial random noise stimulation"[tiab:~0] OR TMS[Title/Abstract] OR "transcranial magnetic stimulation"[tiab:~0] OR TBS[Title/Abstract] OR "theta burst stimulation"[tiab:~0] OR Manual[Title/Abstract] OR "Case Series"[tiab:~0] OR Randomized[Title/Abstract] OR Trial[Title/Abstract] OR Controlled[Title/Abstract] AND Systematic Review[Title/Abstract] OR Meta-Analysis[Title/Abstract])

### Units of Analysis In Included Meta-Analyses

*Supplementary Table 2: Description of all units of analysis among included meta-analyses*

| Study | UOA Description | N | Participants Groups | PUI_Dimension(s) | Treatment(s) | Effect Size | Se Effect Size | Outlier |
|---|---|---|---|---|---|---|---|---|
| **Andrade et al. (2022)** | NA | 15 | Mixed | Internet Addiction | Behavioral | -1.3 | 0.362 | No |
| **Augner et al. (2022)** | PUI Dimension 1 | 14 | Mixed | Internet Addiction | Behavioral | -1.4 | 0.253 | No |
| **Augner et al. (2022)/1** | PUI Dimension 2 | 4 | Mixed | Smartphone | Behavioral | -0.4 | 0.199 | Yes |
| **Chang et al. (2022)** | NA | 29 | Mixed | Internet Addiction Online Gaming | Behavioral + Pharmacological + Neuromodulatory | -1.4 | 0.065 | No |
| **Fendel et al. (2024)** | RCTs | 19 | Mixed | Internet Addiction | Behavioral | -1.7 | 0.245 | No |
| **Fendel et al. (2024)/1** | NRTs | 35 | Mixed | Internet Addiction | Behavioral | -1.7 | 0.161 | No |
| **Goslar et al. (2020)** | IA-Psych-GS | 54 | Mixed | Internet Addiction | Behavioral | -1.5 | 0.11 | No |
| **Goslar et al. (2020)/1** | IA-Psych-FR | 17 | Mixed | Internet Addiction | Behavioral | -1.1 | 0.181 | No |
| **Goslar et al. (2020)/2** | IA-Pharm-GS | 8 | Mixed | Internet Addiction | Pharmacological | -1.1 | 0.146 | No |
| **Goslar et al. (2020)/3** | IA-Pharm-FR | 3 | Mixed | Internet Addiction | Pharmacological | -0.7 | 0.12 | Yes |
| **Goslar et al. (2020)/4** | IA-Comb-GS | 7 | Mixed | Internet Addiction | Behavioral + Pharmacological | -2.5 | 0.415 | Yes |
| **Goslar et al. (2020)/5** | IA-Comb-FR | 2 | Mixed | Internet Addiction | Behavioral + Pharmacological | -2.8 | 0.242 | Yes |
| **Goslar et al. (2020)/6** | SA-Psych-GS | 14 | Mixed | Online Pornography | Behavioral | -1.1 | 0.181 | No |
| **Goslar et al. (2020)/7** | SA-Psych-FR | 6 | Mixed | Online Pornography | Behavioral | -0.8 | 0.145 | Yes |
| **Goslar et al. (2020)/8** | SA-Pharm-GS | 5 | Mixed | Online Pornography | Pharmacological | -1.2 | 0.168 | No |
| **Goslar et al. (2020)/9** | SA-Pharm-FR | 3 | Mixed | Online Pornography | Pharmacological | -0.9 | 0.125 | Yes |
| **Goslar et al. (2020)/10** | CB-Psych-GS | 7 | Mixed | Online Shopping | Behavioral | -1 | 0.128 | No |
| **Goslar et al. (2020)/11** | CB-Psych-FR | 2 | Mixed | Online Shopping | Behavioral | -1 | 0.148 | No |
| **Goslar et al. (2020)/12** | CB-Pharm-GS | 7 | Mixed | Online Shopping | Pharmacological | -1.5 | 0.173 | No |
| **Jiang et al. (2024)** | NA | 66 | Mixed | Internet Addiction | Behavioral + Physical | -2 | 0.143 | Yes |
| **Kim & Noch (2019)** | Pscychological | 4 | Mixed | Internet Addiction | Behavioral | -1.5 | 0.632 | No |
| **Kim & Noch (2019)/1** | Group CBT | 2 | Mixed | Online Gaming | Behavioral | -0.3 | 0.42 | Yes |
| **Li et al. (2023)** | NA | 12 | Adolescents | Smartphone | Physical | -3.1 | 0.41 | Yes |
| **Liu et al. (2017)** | Group Counseling | 30 | Mixed | Internet Addiction | Behavioral | -1.4 | 0.265 | No |
| **Liu et al. (2017)/1** | CBT | 13 | Mixed | Internet Addiction | Behavioral | -1.9 | 0.331 | No |

| Study | UOA Description | N | Participants Groups | PUI_Dimension(s) | Treatment(s) | Effect Size | Se Effect Size | Outlier |
|---|---|---|---|---|---|---|---|---|
| **Liu et al. (2017)/2** | Physical | 9 | Mixed | Internet Addiction | Physical | -1.7 | 0.224 | No |
| **Liu et al. (2019)** | NA | 9 | Younge Adults | Smartphone | Physical | -1.3 | 0.117 | No |
| **Malinauskas & Malinauskiene (2019)** | RCTs | 6 | Adolescents | Internet Addiction<br>Smartphone | Behavioral | -0.7 | 0.204 | Yes |
| **Malinauskas & Malinauskiene (2019)** | RCTs with CBT | 3 | Adolescents | Internet Addiction<br>Smartphone | Behavioral | -0.6 | 0.4 | No |
| **Malinauskas & Malinauskiene (2019)/1** | RCTs with EP | 2 | Adolescents | Internet Addiction<br>Smartphone | Behavioral | -0.8 | 0.319 | No |
| **Stevens et al. (2018)** | NA | 12 | Mixed | Online Gaming | Behavioral | -0.9 | 0.214 | No |
| **Wang et al. (2024)** | NA | 18 | Mixed | Internet Addiction | Behavioral | -1.4 | 0.449 | No |
| **Winkler et al. (2013)** | IA | 16 | Mixed | Internet Addiction | Behavioral + Pharmacological | -1.6 | 0.22 | No |
| **Winkler et al. (2013)** | Time Spent | 16 | Mixed | Internet Addiction | Behavioral + Pharmacological | -0.9 | 0.39 | No |
| **Yan et al. (2024)** | NA | 15 | Young Adults | Internet Addiction | Physical | -1.25 | 0.133 | No |
| **Yeun et al. (2016)** | NA | 37 | Children | Internet Addiction | Behavioral | -1.2 | 0.166 | No |
| **Zhang et al. (2022)** | NA | 24 | Mixed | Internet Addiction<br>Online Gaming<br>Online<br>Pornography | Behavioral + Pharmacological + Neuromodulatory | -1.9 | 0.181 | Yes |
| **Zhang et al. (2024)** | NA | 44 | Young Adults | Internet Addiction | Behavioral + Physical | -1.7 | 0.186 | No |
| **Zhou et al. (2024)** | NA | 89 | Adolescents | Internet Addiction | Behavioral + Physical + Pharmacological + Neuromodulatory | -1.8 | 0.161 | No |
| **Zhu et al. (2023)** | Combined | 57 | Mixed | Internet Addiction<br>Smartphone<br>Online Gaming | Behavioral + Pharmacological + Neuromodulatory | -2.3 | 0.51 | No |
| **Zhu et al. (2023)/1** | EEG Biofeedback | 57 | Mixed | Internet Addiction<br>Smartphone<br>Online Gaming | EEG Biofeedback | -1.6 | 0.34 | No |
| **Zhu et al. (2023)/2** | rTMS | 57 | Mixed | Internet Addiction<br>Smartphone<br>Online Gaming | Neuromodulatory | -1.2 | 0.263 | No |

**General Characteristics of Included Meta-Analyses**

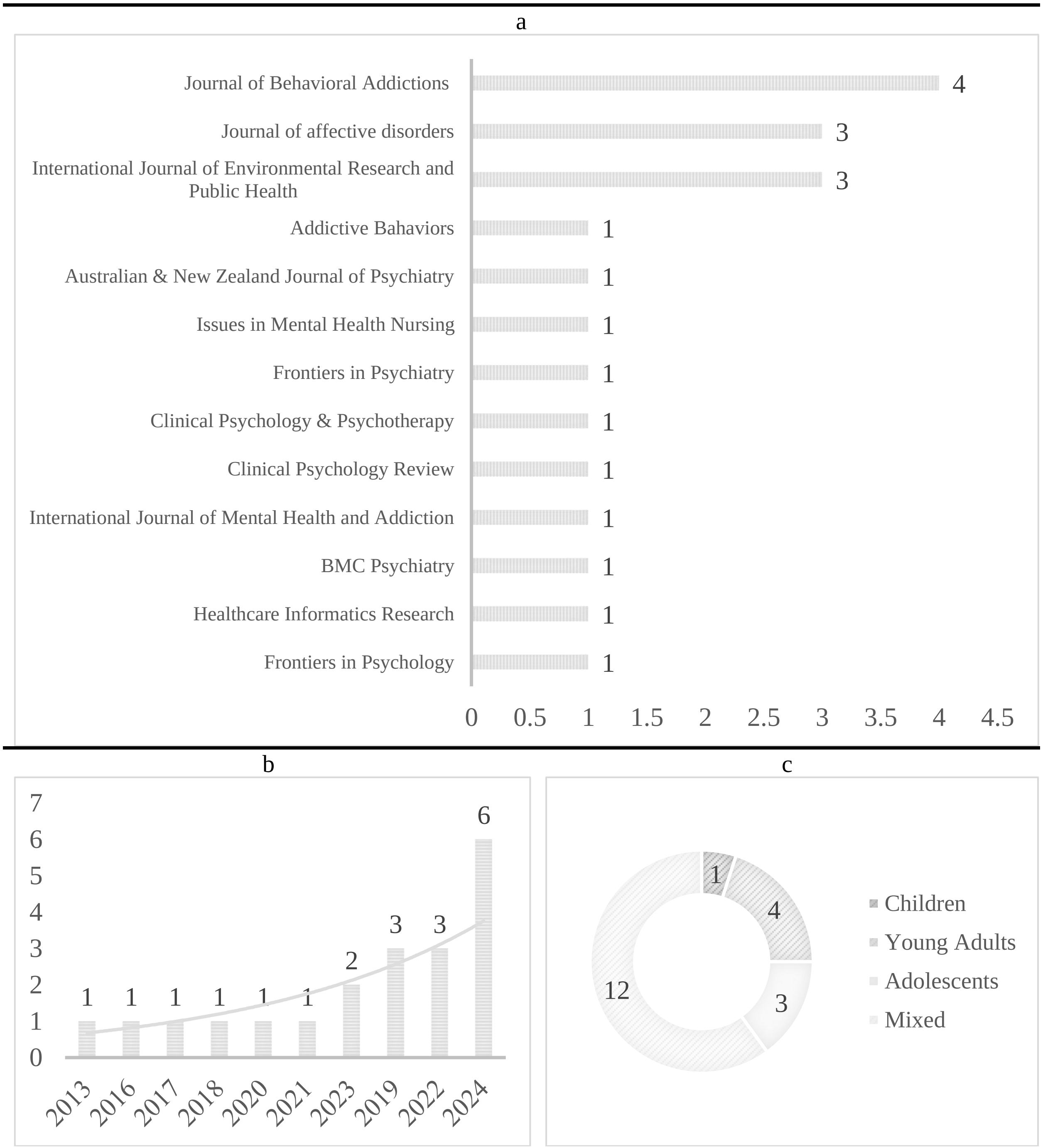


*Supplementary Figure 1: Overview of Publication Trends and Sample Characteristics in Included Meta-Analyses: (Top) Frequency of Meta-Analyses Published in Field-Specific Journals; (Bottom Left) Distribution of Meta-Analyses by Publication Year; (Bottom Right) Age Group Distribution of Participants in the Included Meta-Analyses.*

### Calculating Effect Sizes for Each PUI Dimension and Treatment Category

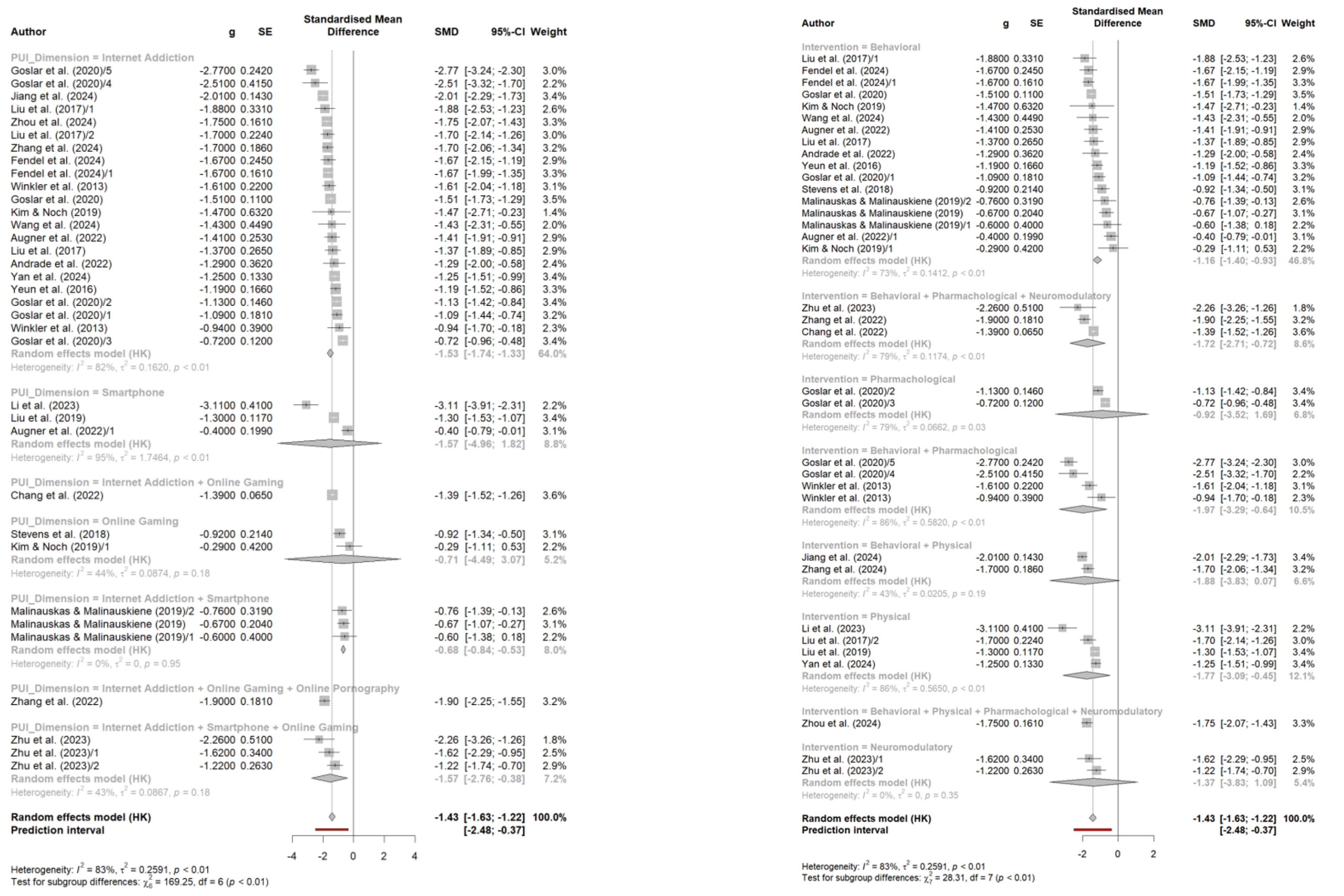


*Supplementary Figure 2: Calculating effect sizes for each PUI dimension (left) and treatment category (right).*

### Evaluating Risk or Bias in Included Meta-analyses with the AMSTAR Checklist

*Supplementary Table 3: Evaluating Risk or Bias in Included Meta-Analyses with AMSTAR Checklist*

| | Andrade et al. (2022) | Augner et al. (2022) | Chang et al. (2022) | Fendel et al. (2024) | Goslar et al. (2020) | Jiang et al. (2024) | Kim & Noch (2019) | Li et al. (2023) | Liu et al. (2017) | Liu et al. (2019) | Malinauskas & Malinauskiene (2019) | Stevens et al. (2018) | Wang et al. (2024) | Winkler et al. (2013) | Yan et al. (2024) | Yeun et al. (2016) | Zhang et al. (2022) | Zhang et al. (2024) | Zhou et al. (2024) | Zhu et al. (2023) |
|---|---|---|---|---|---|---|---|---|---|---|---|---|---|---|---|---|---|---|---|---|
| **1. Did the research questions and inclusion criteria for The review includes the components of PICO.** | Yes | Yes | Yes | Yes | Yes | Yes | Yes | Yes | Yes | Yes | Yes | Yes | Yes | Yes | Yes | Yes | Yes | Yes | Yes | Yes |
| **2. Did the report of the review contain an explicit statement that the review methods were established prior to the conduct of the review, and did the report justify any significant deviations from the protocol?** | Yes | Partial Yes | Partial Yes | Yes | Partial Yes | Partial Yes | Partial Yes | Partial Yes | Partial Yes | Partial Yes | Partial Yes | Partial Yes | Partial Yes | Partial Yes | Yes | Partial Yes | Yes | Yes | Yes | Partial Yes |
| **3. Did the review authors explain their selection of the study designs for inclusion in the review?** | No | No | No | No | No | No | No | No | No | No | No | No | No | No | Yes | No | No | No | No | No |
| **4. Did the review authors use a comprehensive literature search strategy?** | Yes | Yes | Yes | Yes | Yes | Yes | Yes | Yes | Yes | Yes | Yes | Yes | Yes | Yes | Yes | Yes | Yes | Yes | Yes | Yes |
| **5. Did the review authors perform study selection in duplicate?** | Yes | Yes | Yes | Yes | Yes | Yes | Yes | Yes | Yes | Yes | Yes | Yes | Yes | Yes | Yes | Yes | Yes | Yes | Yes | Yes |
| **6. Did the review authors perform data extraction in duplicate?** | Yes | Yes | Yes | Yes | Yes | Yes | Yes | Yes | Yes | Yes | Yes | Yes | Yes | Yes | Yes | Yes | Yes | Yes | Yes | Yes |
| **7. Did the review authors provide a list of excluded studies and justify the exclusions?** | No | No | No | No | No | No | No | No | No | No | No | No | No | No | No | No | No | No | No | No |
| **8. Did the review authors describe the included studies in adequate detail?** | Yes | Yes | Yes | Yes | Yes | Yes | Yes | Yes | Yes | Yes | Yes | Yes | Yes | Yes | Yes | Yes | Yes | Yes | Yes | Yes |

| | Andrade et al. (2022) | Augner et al. (2022) | Chang et al. (2022) | Fendel et al. (2024) | Goslar et al. (2020) | Jiang et al. (2024) | Kim & Noch (2019) | Li et al. (2023) | Liu et al. (2017) | Liu et al. (2019) | Malinauskas & Malinauskiene (2019) | Stevens et al. (2018) | Wang et al. (2024) | Winkler et al. (2013) | Yan et al. (2024) | Yeun et al. (2016) | Zhang et al. (2022) | Zhang et al. (2024) | Zhou et al. (2024) | Zhu et al. (2023) |
|---|---|---|---|---|---|---|---|---|---|---|---|---|---|---|---|---|---|---|---|---|
| **9. Did the review authors use a satisfactory technique for assessing the risk of bias (RoB) in individual studies that were included in the review?** | Yes | No | No | Yes | Yes | Yes | No | Yes | Yes | Yes | Yes | No | Yes | No | Yes | Yes | Yes | Yes | Yes | Yes |
| **10. Did the review authors report on the sources of funding for the studies included in the review?** | No | No | No | No | No | No | No | No | No | No | No | No | No | No | No | No | No | No | No | No |
| **11. If meta-analysis was performed, did the review authors use appropriate methods for statistical combination of results?** | Yes | Yes | Yes | Yes | Yes | Yes | Yes | Yes | Yes | Yes | Yes | No | Yes | Yes | Yes | Yes | Yes | Yes | Yes | Yes |
| **12. If meta-analysis was performed, did the review authors assess the potential impact of RoB in individual studies on the results of the meta-analysis or other evidence synthesis?** | Yes | No | No | Yes | Yes | Yes | No | Yes | Yes | Yes | No | Yes | Yes | No | Yes | Yes | Yes | Yes | Yes | Yes |
| **13. Did the review authors account for RoB in individual studies when interpreting/ discussing the results of the review?** | Yes | No | No | Yes | Yes | Yes | No | Yes | Yes | Yes | Yes | No | Yes | No | Yes | Yes | Yes | Yes | Yes | Yes |
| **14. Did the review authors provide a satisfactory explanation for, and discussion of, any heterogeneity observed in the results of the review?** | Yes | Yes | Yes | Yes | Yes | Yes | Yes | Yes | Yes | Yes | Yes | Yes | Yes | Yes | Yes | Yes | Yes | Yes | Yes | Yes |
| **15. If they performed quantitative synthesis, did the review authors carry out an adequate investigation of publication bias (small study bias) and discuss its likely impact on the results of the review?** | Yes | Yes | Yes | Yes | Yes | Yes | No | Yes | Yes | Yes | Yes | Yes | Yes | Yes | Yes | Yes | Yes | Yes | Yes | Yes |

| | Andrade et al. (2022) | Augner et al. (2022) | Chang et al. (2022) | Fendel et al. (2024) | Goslar et al. (2020) | Jiang et al. (2024) | Kim & Noch (2019) | Li et al. (2023) | Liu et al. (2017) | Liu et al. (2019) | Malinauskas & Malinauskiene (2019) | Stevens et al. (2018) | Wang et al. (2024) | Winkler et al. (2013) | Yan et al. (2024) | Yeun et al. (2016) | Zhang et al. (2022) | Zhang et al. (2024) | Zhou et al. (2024) | Zhu et al. (2023) |
|---|---|---|---|---|---|---|---|---|---|---|---|---|---|---|---|---|---|---|---|---|
| **16. Did the review authors report any potential sources of conflict of interest, including any funding they received for conducting the review?** | Yes | Yes | Yes | Yes | Yes | Yes | Yes | Yes | Yes | Yes | Yes | Yes | Yes | No | No | Yes | Yes | Yes | Yes | Yes |
| **Overall Risk of Bias** | **Low** | **Low** | **Critically Low** | **Low** | **Low** | **Low** | **Critically Low** | **Low** | **Low** | **Low** | **Critically Low** | **Critically Low** | **Low** | **Critically Low** | | **Low** | **Low** | **Low** | **Low** | **Low** |

**Funnel Plot For Publication Bias in Meta-Analyses**

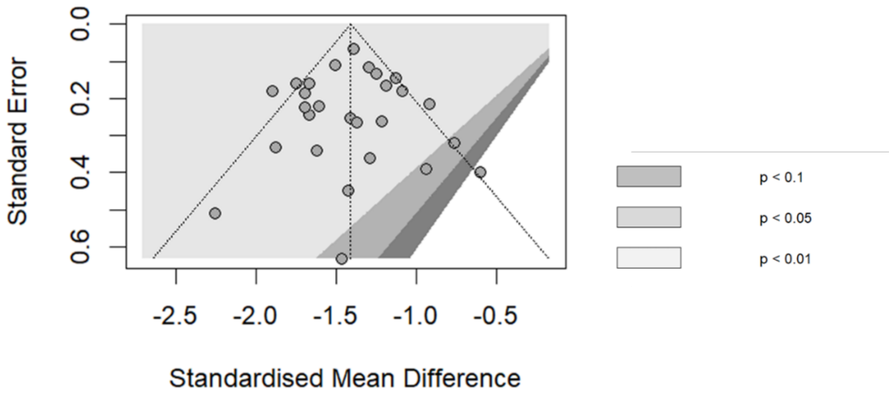


*Supplementary Figure 3: Funnel Plot for Included Meta-Analyses after removing outliers*

**Subgroup Analysis in Meta-Analyses**

*Supplementary Table 4: Subgroup Analysis for Participant Groups in Meta-Analysis*

| Parameter | Contrast | K | SMD | 95%-CI | tau^2 | Q | I^2 | Test for differences |
|---|---|---|---|---|---|---|---|---|
| Participants Groups | Mixed | 20 | -1.4475 | [-1.5870; -1.3081] | 0.0375 | 33.62 | 43.5% | Q = 2.79<br>df = 3<br>P-value = 0.4258 |
| | Young Adults | 3 | -1.3832 | [-1.9471; -0.8193] | 0.0240 | 4.28 | 53.3% | |
| | Adolescents | 3 | -1.0994 | [-2.6909; 0.4920] | 0.3499 | 12.66 | 84.2% | |
| | Children | 1 | -1.1900 | [-1.5154; -0.8646] | - | 0.00 | - | |

**Meta-regression Analysis in Meta-Analyses**

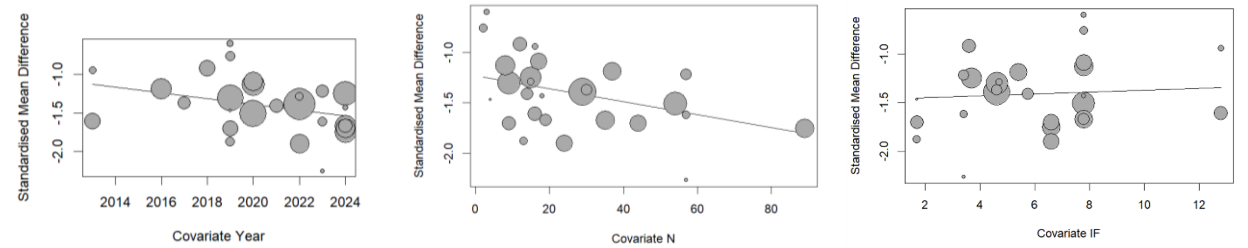


*Supplementary Figure 4: Meta regression Bubble Diagram of the Effect of Publishing Year (left), Number of Participants (middle), and Journals' Impact Factor (right) on the Effect Sizes in the Included Meta-Analyses.*

**PUI Dimenstions in RCTs**

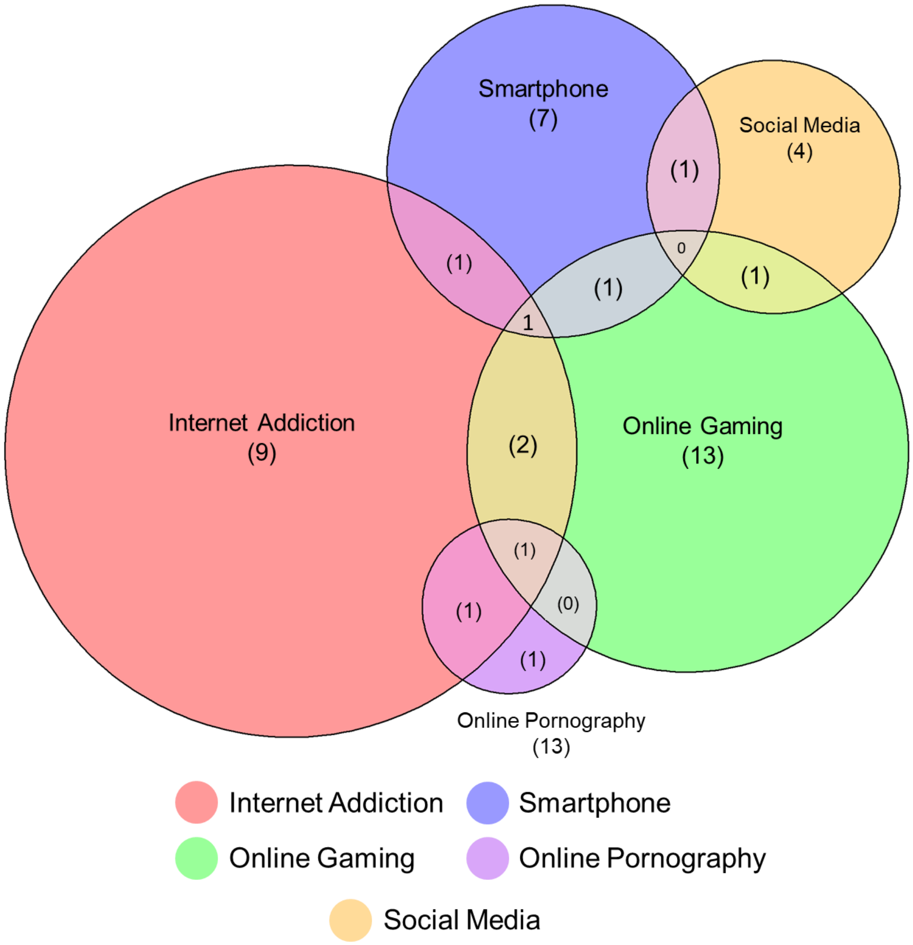


*Supplementary Figure 5 the distribution of PUI dimensions addressed across the included RCTs, highlighting patterns in topic representation and overlap.*

### Calculating Effect Size Based on PUI Dimensions and Treatment Categories

a

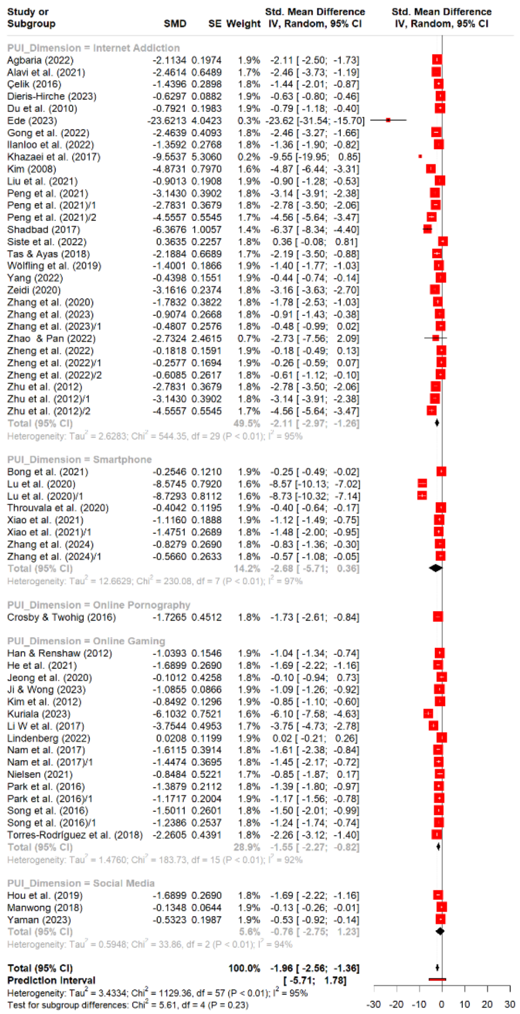


b

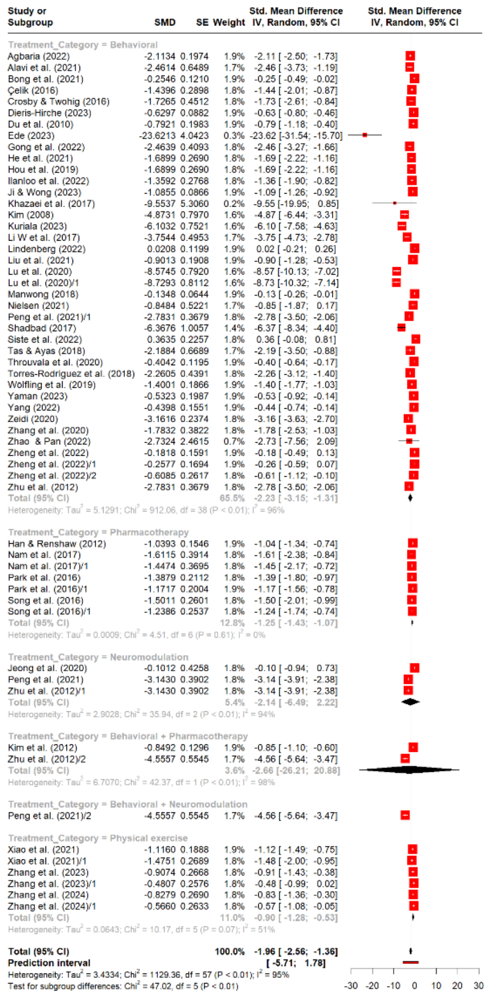


*Supplementary Figure 6 Results of Effect Size Calculation on the PUI Dimensions and Treatment Categories (N=58 units of analysis)*

| Study or Subgroup | SMD | SE | Weight | Std. Mean Difference IV, Random, 95% CI |
|---|---|---|---|---|
| subgroup = Internet Addiction.Behavioral | | | | |
| Agbaria (2022) | -2.1134 | 0.1974 | 1.9% | -2.11 [ -2.50; -1.73] |
| Alavi et al. (2021) | -2.4614 | 0.6489 | 1.7% | -2.46 [ -3.73; -1.19] |
| Çelik (2016) | -1.4396 | 0.2898 | 1.8% | -1.44 [ -2.01; -0.87] |
| Dieris-Hirche (2023) | -0.6297 | 0.0882 | 1.9% | -0.63 [ -0.80; -0.46] |
| Du et al. (2010) | -0.7921 | 0.1983 | 1.9% | -0.79 [ -1.18; -0.40] |
| Ede (2023) | -23.6213 | 4.0423 | 0.3% | -23.62 [-31.54; -15.70] |
| Gong et al. (2022) | -2.4639 | 0.4093 | 1.8% | -2.46 [ -3.27; -1.66] |
| Ilanloo et al. (2022) | -1.3592 | 0.2768 | 1.8% | -1.36 [ -1.90; -0.82] |
| Khazaei et al. (2017) | -9.5537 | 5.3060 | 0.2% | -9.55 [-19.95; 0.85] |
| Kim (2008) | -4.8731 | 0.7970 | 1.6% | -4.87 [ -6.44; -3.31] |
| Liu et al. (2021) | -0.9013 | 0.1908 | 1.9% | -0.90 [ -1.28; -0.53] |
| Peng et al. (2021)/1 | -2.7831 | 0.3679 | 1.8% | -2.78 [ -3.50; -2.06] |
| Shadbad (2017) | -6.3676 | 1.0057 | 1.4% | -6.37 [ -8.34; -4.40] |
| Siste et al. (2022) | 0.3635 | 0.2257 | 1.8% | 0.36 [ -0.08; 0.81] |
| Tas & Ayas (2018) | -2.1884 | 0.6689 | 1.7% | -2.19 [ -3.50; -0.88] |
| Wölfling et al. (2019) | -1.4001 | 0.1866 | 1.9% | -1.40 [ -1.77; -1.03] |
| Yang (2022) | -0.4398 | 0.1551 | 1.9% | -0.44 [ -0.74; -0.14] |
| Zeidi (2020) | -3.1616 | 0.2374 | 1.8% | -3.16 [ -3.63; -2.70] |
| Zhang et al. (2020) | -1.7832 | 0.3822 | 1.8% | -1.78 [ -2.53; -1.03] |
| Zhao & Pan (2022) | -2.7324 | 2.4615 | 0.7% | -2.73 [ -7.56; 2.09] |
| Zheng et al. (2022) | -0.1818 | 0.1591 | 1.9% | -0.18 [ -0.49; 0.13] |
| Zheng et al. (2022)/1 | -0.2577 | 0.1694 | 1.9% | -0.26 [ -0.59; 0.07] |
| Zheng et al. (2022)/2 | -0.6085 | 0.2617 | 1.8% | -0.61 [ -1.12; -0.10] |
| Zhu et al. (2012) | -2.7831 | 0.3679 | 1.8% | -2.78 [ -3.50; -2.06] |
| Total (95% CI) | | | 38.8% | -1.91 [ -2.91; -0.92] |
| Heterogeneity: $Tau^2 = 2.3535$; $Chi^2 = 397.49$, df = 23 ($P < 0.01$); $I^2 = 94\%$ | | | | |
| subgroup = Online Gaming.Behavioral | | | | |
| He et al. (2021) | -1.6899 | 0.2690 | 1.8% | -1.69 [ -2.22; -1.16] |
| Ji & Wong (2023) | -1.0855 | 0.0866 | 1.9% | -1.09 [ -1.26; -0.92] |
| Kuriala (2023) | -6.1032 | 0.7521 | 1.6% | -6.10 [ -7.58; -4.63] |
| Li W et al. (2017) | -3.7544 | 0.4953 | 1.7% | -3.75 [ -4.73; -2.78] |
| Lindenberg (2022) | 0.0208 | 0.1199 | 1.9% | 0.02 [ -0.21; 0.26] |
| Nielsen (2021) | -0.8484 | 0.5221 | 1.7% | -0.85 [ -1.87; 0.17] |
| Torres-Rodríguez et al. (2018) | -2.2605 | 0.4391 | 1.8% | -2.26 [ -3.12; -1.40] |
| Total (95% CI) | | | 12.4% | -2.17 [ -4.05; -0.29] |
| Heterogeneity: $Tau^2 = 3.7813$; $Chi^2 = 162.85$, df = 6 ($P < 0.01$); $I^2 = 96\%$ | | | | |
| subgroup = Online Pornography.Behavioral | | | | |
| Crosby & Twohig (2016) | -1.7265 | 0.4512 | 1.8% | -1.73 [ -2.61; -0.84] |
| subgroup = Smartphone.Behavioral | | | | |
| Bong et al. (2021) | -0.2546 | 0.1210 | 1.9% | -0.25 [ -0.49; -0.02] |
| Lu et al. (2020) | -8.5745 | 0.7920 | 1.6% | -8.57 [-10.13; -7.02] |
| Lu et al. (2020)/1 | -8.7293 | 0.8112 | 1.6% | -8.73 [-10.32; -7.14] |
| Throuvala et al. (2020) | -0.4042 | 0.1195 | 1.9% | -0.40 [ -0.64; -0.17] |
| Total (95% CI) | | | 6.9% | -4.43 [-12.08; 3.21] |
| Heterogeneity: $Tau^2 = 22.7637$; $Chi^2 = 211.59$, df = 3 ($P < 0.01$); $I^2 = 99\%$ | | | | |
| subgroup = Social Media.Behavioral | | | | |
| Hou et al. (2019) | -1.6899 | 0.2690 | 1.8% | -1.69 [ -2.22; -1.16] |
| Manwong (2018) | -0.1348 | 0.0644 | 1.9% | -0.13 [ -0.26; -0.01] |
| Yaman (2023) | -0.5323 | 0.1987 | 1.9% | -0.53 [ -0.92; -0.14] |
| Total (95% CI) | | | 5.6% | -0.76 [ -2.75; 1.23] |
| Heterogeneity: $Tau^2 = 0.5948$; $Chi^2 = 33.86$, df = 2 ($P < 0.01$); $I^2 = 94\%$ | | | | |
| subgroup = Internet Addiction.Behavioral + Neuromodulation | | | | |
| Peng et al. (2021)/2 | -4.5557 | 0.5545 | 1.7% | -4.56 [ -5.64; -3.47] |
| subgroup = Internet Addiction.Behavioral + Pharmacotherapy | | | | |
| Zhu et al. (2012)/2 | -4.5557 | 0.5545 | 1.7% | -4.56 [ -5.64; -3.47] |
| subgroup = Online Gaming.Behavioral + Pharmacotherapy | | | | |
| Kim et al. (2012) | -0.8492 | 0.1296 | 1.9% | -0.85 [ -1.10; -0.60] |
| subgroup = Internet Addiction.Neuromodulation | | | | |
| Peng et al. (2021) | -3.1430 | 0.3902 | 1.8% | -3.14 [ -3.91; -2.38] |
| Zhu et al. (2012)/1 | -3.1430 | 0.3902 | 1.8% | -3.14 [ -3.91; -2.38] |
| Total (95% CI) | | | 3.6% | -3.14 [ -3.14; -3.14] |
| Heterogeneity: $Tau^2 = 0$; $Chi^2 = 0$, df = 1 ($P = 1.00$); $I^2 = 0\%$ | | | | |
| subgroup = Online Gaming.Neuromodulation | | | | |
| Jeong et al. (2020) | -0.1012 | 0.4258 | 1.8% | -0.10 [ -0.94; 0.73] |
| subgroup = Online Gaming.Pharmacotherapy | | | | |
| Han & Renshaw (2012) | -1.0393 | 0.1546 | 1.9% | -1.04 [ -1.34; -0.74] |
| Nam et al. (2017) | -1.6115 | 0.3914 | 1.8% | -1.61 [ -2.38; -0.84] |
| Nam et al. (2017)/1 | -1.4474 | 0.3695 | 1.8% | -1.45 [ -2.17; -0.72] |
| Park et al. (2016) | -1.3879 | 0.2112 | 1.8% | -1.39 [ -1.80; -0.97] |
| Park et al. (2016)/1 | -1.1717 | 0.2004 | 1.9% | -1.17 [ -1.56; -0.78] |
| Song et al. (2016) | -1.5011 | 0.2601 | 1.8% | -1.50 [ -2.01; -0.99] |
| Song et al. (2016)/1 | -1.2386 | 0.2537 | 1.8% | -1.24 [ -1.74; -0.74] |
| Total (95% CI) | | | 12.8% | -1.25 [ -1.43; -1.07] |
| Heterogeneity: $Tau^2 = 0.0009$; $Chi^2 = 4.51$, df = 6 ($P = 0.61$); $I^2 = 0\%$ | | | | |
| subgroup = Internet Addiction.Physical exercise | | | | |
| Zhang et al. (2023) | -0.9074 | 0.2668 | 1.8% | -0.91 [ -1.43; -0.38] |
| Zhang et al. (2023)/1 | -0.4807 | 0.2576 | 1.8% | -0.48 [ -0.99; 0.02] |
| Total (95% CI) | | | 3.7% | -0.69 [ -3.40; 2.02] |
| Heterogeneity: $Tau^2 = 0.0223$; $Chi^2 = 1.32$, df = 1 ($P = 0.25$); $I^2 = 24\%$ | | | | |
| subgroup = Smartphone.Physical exercise | | | | |
| Xiao et al. (2021) | -1.1160 | 0.1888 | 1.9% | -1.12 [ -1.49; -0.75] |
| Xiao et al. (2021)/1 | -1.4751 | 0.2689 | 1.8% | -1.48 [ -2.00; -0.95] |
| Zhang et al. (2024) | -0.8279 | 0.2690 | 1.8% | -0.83 [ -1.36; -0.30] |
| Zhang et al. (2024)/1 | -0.5660 | 0.2633 | 1.8% | -0.57 [ -1.08; -0.05] |
| Total (95% CI) | | | 7.4% | -1.00 [ -1.60; -0.40] |
| Heterogeneity: $Tau^2 = 0.0740$; $Chi^2 = 6.6$, df = 3 ($P = 0.09$); $I^2 = 55\%$ | | | | |
| **Total (95% CI)** | | | **100.0%** | **-1.96 [ -2.56; -1.36]** |
| **Prediction interval** | | | | **[ -5.71; 1.78]** |
| Heterogeneity: $Tau^2 = 3.4334$; $Chi^2 = 1129.36$, df = 57 ($P < 0.01$); $I^2 = 95\%$ | | | | |
| Test for subgroup differences: $Chi^2 = 102.21$, df = 12 ($P < 0.01$) | | | | |

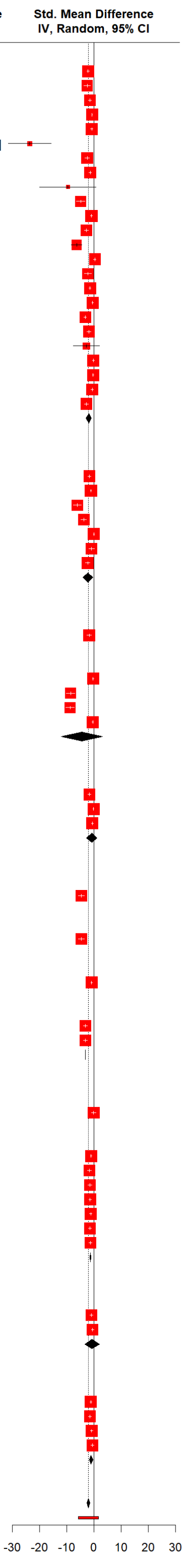


*Supplementary Figure 7: Analysis of effect sizes on the interaction of PUI dimensions and treatment approaches*

**Funnel Plot For Publication Bias in RCTs**

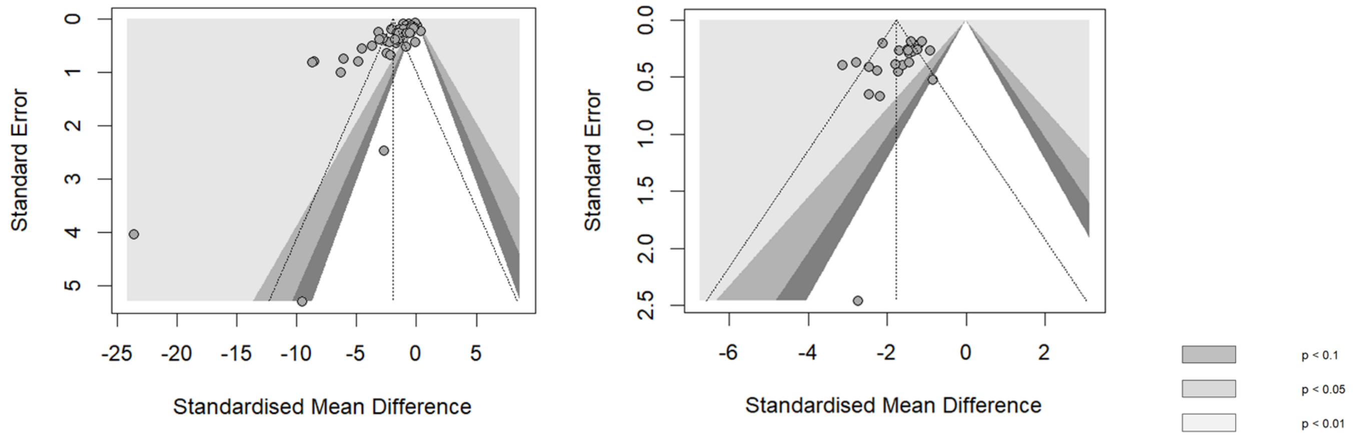


*Supplementary Figure 8: Funnel Plot for RCTs before (left) and after (right) outliers removal*

### Risk of Bias in RCTs with RoB 2 Tool

| Sudy | D1 | D2 | D3 | D4 | D5 | Overall | Sudy | D1 | D2 | D3 | D4 | D5 | Overall |
|---|---|---|---|---|---|---|---|---|---|---|---|---|---|
| Agbaria (2022) | ✓ | ✓ | ✓ | ✓ | ✓ | ✓ | Lu et al. (2020) | ✓ | ! | ✓ | ! | ! | ✗ |
| Alavi et al. (2021) | ! | ✓ | ✓ | ✓ | ! | ! | Manwong (2018) | ! | ! | ✓ | ! | ✓ | ✗ |
| Bong et al. (2021) | ! | ✓ | ✓ | ✓ | ! | ! | Nam et al. (2017) | ! | ✓ | ✓ | ✓ | ! | ! |
| Çelik (2016) | ✗ | ! | ✓ | ✗ | ! | ✗ | Nielsen (2021) | ✓ | ! | ✓ | ✓ | ! | ! |
| Crosby & Twohig (2016) | ✗ | ✓ | ✓ | ! | ! | ✗ | Park et al. (2016) | ! | ✓ | ✓ | ! | ! | ✗ |
| Dieris-Hirche (2023) | ! | ✓ | ✓ | ✓ | ✓ | ! | Peng et al. (2021) | ✓ | ✓ | ✓ | ✓ | ✓ | ✓ |
| Du et al. (2010) | ! | ✓ | ✓ | ✓ | ! | ! | Shadbad (2017) | ! | ✓ | ✓ | ! | ! | ✗ |
| Ede (2023) | ✓ | ! | ✓ | ! | ! | ✗ | Siste et al. (2022) | ! | ✓ | ✓ | ✓ | ✓ | ! |
| Gong et al. (2022) | ✗ | ! | ✓ | ! | ! | ✗ | Song et al. (2016) | ! | ✓ | ✓ | ✗ | ! | ✗ |
| Han & Renshaw (2012) | ! | ✓ | ✓ | ✓ | ! | ! | Tas & Ayas (2018) | ! | ✓ | ✓ | ! | ! | ✗ |
| He et al. (2021) | ! | ✓ | ✓ | ✓ | ! | ! | Torres-Rodríguez et al. (2018) | ! | ✓ | ✓ | ✓ | ! | ! |
| Hou et al. (2019) | ! | ! | ✓ | ! | ! | ✗ | Wölfling et al. (2019) | ! | ! | ✓ | ! | ✓ | ✗ |
| Ilanloo et al. (2022) | ! | ! | ✓ | ! | ! | ✗ | Xiao et al. (2021) | ! | ✓ | ✓ | ! | ! | ✗ |
| Jeong et al. (2020) | ! | ✓ | ✓ | ! | ! | ✗ | Yaman (2023) | ! | ✓ | ✓ | ! | ✓ | ! |
| Ji & Wong (2023) | ! | ✓ | ✓ | ✓ | ! | ! | Yang (2022) | ! | ! | ✓ | ! | ! | ✗ |
| Khazaei et al. (2017) | ! | ✓ | ✓ | ✗ | ! | ✗ | Zeidi (2020) | ! | ! | ✓ | ! | ! | ✗ |
| Kim (2008) | ! | ✓ | ✓ | ! | ! | ✗ | Zhang et al. (2020) | ! | ! | ✓ | ! | ! | ✗ |
| Kim et al. (2012) | ! | ! | ✓ | ✗ | ! | ✗ | Zhang et al. (2023) | ✓ | ✓ | ✓ | ✓ | ✓ | ✓ |
| Kuriala (2023) | ! | ! | ✗ | ! | ! | ✗ | Zhang et al. (2024) | ✓ | ✓ | ✓ | ✓ | ✓ | ✓ |
| Li W et al. (2017) | ! | ✓ | ✓ | ! | ! | ✗ | Zhao & Pan (2022) | ! | ✓ | ✓ | ! | ! | ✗ |
| Lindenberg (2022) | ! | ✓ | ✓ | ! | ✓ | ! | Zheng et al. (2022) | ! | ! | ✓ | ! | ! | ✗ |
| Liu et al. (2021) | ! | ✓ | ✓ | ✓ | ! | ! | Zhu et al. (2012) | ! | ✓ | ✓ | ! | ! | ✗ |

*Supplementary Figure 9 Supplementary Figure 7 Results of Risk of Bias Evaluation in RCTs with RoB2: Low (✓), Some Concerns (!), and High(✗)*

### Subgroup Analysis in RCTs

*Supplementary Table 5: Summary of Subgroup Analysis for RCTs after outlier removal*

| Parameter | Contrast | K | SMD 95%-CI | $tau^2$ | Q | $I^2$ | Test for differences |
|---|---|---|---|---|---|---|---|
| All units of analysis | - | 26 | -1.7660 [-2.0310; -1.5010] | 0.3025 | 94.28 | 73.5% | |
| Cultural Background | Non-East Asian | 9 | -1.6845 [-2.0396; -1.3294] | 0.0869 | 15.42 | 48.1% | Q = 0.28, df = 1, p-value = 0.5944 |
| | Eastern Asian | 17 | -1.8103 [-2.1900; -1.4306] | 0.4294 | 78.37 | 79.6% | |
| Control Conditions | Sham Controlled | 3 | -2.1463 [-2.4705; -1.8221] | 0 | 0.32 | 0.0% | Q = 35.19, df = 1, **p-value = < 0.0001** |
| | No Treatment | 13 | -1.4295 [-1.6373; -1.2218] | 0.0206 | 17.17 | 30.1% | |
| | Active Controlled | 10 | -2.0491 [-2.6622; -1.4359] | 0.5943 | 57.11 | 84.2% | |
| Contrast | Symptoms | 25 | -1.7695 [-2.0454; -1.4935] | 0.3186 | 94.22 | 74.5% | Q = 0.01, df = 1, p-value = 0.9273 |
| | Consumption | 1 | -1.7265 [-2.6108; -0.8422] | -- | 0.00 | -- | |
| Participants Groups | Adolescents | 10 | -1.6485 [-2.0251; -1.2720] | 0.1530 | 23.18 | 61.2% | Q = 8.46, df = 1, **p-value = 0.0146** |
| | Young Adults | 8 | -1.3752 [-1.6731; -1.0773] | 0.0292 | 10.85 | 35.5% | |
| | Adults | 8 | -2.2372 [-2.8895; -1.5849] | 0.4860 | 40.12 | 82.6% | |
| PUI Dimensions | Internet Addiction | 14 | -2.1050 [-2.5341; -1.6759] | 0.4469 | 63.38 | 79.5% | Q = 13.83, df = 4, **p-value = 0.0078** |
| | Online Pornography | 1 | -1.7265 [-2.6108; -0.8422] | - | 0.00 | - | |
| | Social Media | 1 | -1.6899 [-2.2172; -1.1626] | | 0.00 | | |
| | Online Gaming | 8 | -1.3721 [-1.6089; -1.1353] | <0.0001 | 7.04 | 0.6% | |
| | Smartphone | 2 | -1.2445 [-3.4318; 0.9428] | 0.0105 | 1.19 | 16.3% | |
| Treatment Category | Behavioral | 15 | -1.9125 [-2.2241; -1.6009] | 0.1807 | 34.10 | 58.9% | Q = 16.19, df = 3, **p-value = 0.0010** |
| | Pharmacotherapy | 6 | -1.3427 [-1.5068; -1.1786] | - | 1.87 | 0.0% | |
| | Neuromodulation | 2 | -3.1430 [-3.1430; -3.1430] | - | 0.00 | 0.0% | |
| | Physical exercise | 3 | -1.1524 [-1.7721; -0.5326] | <0.0001 | 2.32 | 13.8% | |

| **Parameter** | **Contrast** | **K** | **SMD 95%-CI** | **$tau^2$** | **Q** | **$I^2$** | **Test for differences** |
|---|---|---|---|---|---|---|---|
| All units of analysis | - | 26 | -1.7660 [-2.0310; -1.5010] | 0.3025 | 94.28 | 73.5% | |
| Risk of Bias | Low | 4 | -2.2080 [-3.7707; -0.6453] | 0.8654 | 30.20 | 90.1% | Q = 3.12, df = 2, p-value = 0.2099 |
| | Some Concerns | 7 | -1.4495 [-1.8456; -1.0534] | 0.0304 | 9.23 | 35.0% | |
| | High | 15 | -1.7381 [-2.0682; -1.4080] | 0.2355 | 42.89 | 67.4% | |

### Meta Regression in RCTs

*Supplementary Table 6: Summary of Meta-regression Analysis in RCTs*

| **Parameter** | Publication Year | Sample Size | Frequency | Number of Days | Number of Sessions |
|---|---|---|---|---|---|
| **$tau^2$** | 0.3013 (SE = 0.1216) | 0.3170 (SE = 0.1265) | 0.2403 (SE = 0.1025) | 0.2981 (SE = 0.1211) | 0.3279 (SE = 0.1554) |
| **$I^2$** | 77.03% | 77.88% | 72.68% | 77.57% | 76.13% |
| **$H^2$** | 4.35 | 4.52 | 3.66 | 4.46 | 4.19 |
| **$R^2$** | 0.41% | 0.00% | 20.58% | 3.71% | 14.07% |
| **F** | 1.2144 | 0.3802 | 4.6251 | 1.6841 | 3.6557 |
| **p-value** | 0.2814 | 0.5433 | 0.0418 | 0.2072 | 0.0719 |
